# Common and rare variant genetic contributions in African Americans with autism

**DOI:** 10.1101/2025.11.18.25340532

**Authors:** Matilde Cirnigliaro, Jennifer K. Lowe, Alexander O. Flynn-Carroll, Michi E. Kumagai, David S. Gibson, Jamal B. Williams, Jack M. Fu, Shan Dong, Kangcheng Hou, Vamsee Pillalamarri, Anna M. Abbacchi, Amanda C. Gulsrud, Janet Miller, Yi Zhang, Erin T. Graham, Elizabeth O. Akinyemi, Marshel F. Adams, Amaris N. Clay, Stephanie A. Arteaga, Hailey Choi, Ryan M. Kochis, Jorge E. Peña-Velasco, Jackson N. Hoekstra, Aaron D. Besterman, Sunil Mehta, Tarik Hadzic, Rujuta B. Wilson, Tashalee R. Brown, Leanna M. Hernandez, Natasha Marrus, Sophie Molholm, Cheryl Klaiman, Rita M. Cantor, Michael E. Talkowski, Stephan J. Sanders, Dan E. Arking, Bogdan Pasaniuc, Ami Klin, John N. Constantino, Genetics of Neurodevelopment in African Americans (GENAA) Consortium, Daniel H. Geschwind

## Abstract

The absence of non-European cohorts in genetic studies of neurodevelopmental and neuropsychiatric disorders severely limits the understanding of their full genetic architecture and undermines implementation of precision medicine. Here, we directly addressed this issue by recruiting African Americans (AfrAms) with autism spectrum disorder (ASD) and analyzing their rare and common genetic variation. We performed both global and local ancestry analyses to characterize the complex patterns of admixture at the individual level and compare genetic factors between European (EUR) and African (AFR) genetically inferred ancestries (GIAs) across multiple cohorts in a total of 38,483 autistic individuals. We showed consistent common variant genetic effect sizes for ASD in EUR and AFR GIAs through genome-wide association studies. We demonstrated the limited transferability of EUR-derived polygenic scores (PGSs) based on polygenic transmission disequilibrium and ancestry partial PGS analysis. We found significant autism association for high-impact rare copy number variants in both GIAs. We identified a set of candidate ASD loci based on rare deletions observed in AFR GIA carriers, including *SMC2*, *DMTN*, *SORCS1*, and *ROGDI*, and detected a signal for *de novo* missense variants of predicted low impact in AFR GIA individuals. Finally, we uncovered significant depletion of AFR GIA autistic carriers of rare variants in known associated genes found in EUR cohort studies. These findings are the first to detail common and rare variant genetic contributions to ASD in AfrAms and demonstrate that their involvement in neurodevelopmental and neuropsychiatric disorders’ genomic research is essential to advance discovery.

## Introduction

In the United States, 1 in 31 children receive an autism spectrum disorder (ASD) diagnosis^1^, with historically underserved groups such as girls and non-White children^2–5^ steadily showing increased access to ASD evaluation and identification^1,6^. Over the last decade, the understanding of ASD genetic architecture has advanced significantly^7–17^. Twin studies^18^ and other family-design studies^19–22^ have provided estimates of narrow-sense heritability for ASD of 64-91% and 69-85%, respectively. Common variants of small additive effects represent the main genetic factors contributing to ASD narrow-sense heritability, explaining 12-65% of ASD phenotypic variance based on estimates from different study designs^7,9,15,23,24^, and several specific loci have been identified through genome-wide association studies (GWASs), which account for ∼2% of ASD variance^9,14^. In parallel, analyses of high-impact rare inherited and *de novo* variants through whole-genome and whole-exome sequencing (WES) have led to the discovery of more than 200 high-confidence ASD-associated genes^11–13^, and have shown that rare variation contributes to only ∼2-6% of ASD variance^7,11,14^. However, these important findings resulted from ASD studies analyzing individuals of primarily European (EUR) genetic ancestries, consistent with the vast majority of current human genetic research participants^25–27^, but raising questions about their transferability to other populations. Moreover, with a large proportion of heritability for ASD still hidden or unaccounted for, many rare and common genetic contributing factors have yet to be uncovered, which emphasizes the need for larger sample sizes and more heterogeneous study designs. Indeed, recent work has highlighted the role of rare inherited variation in ASD, which had been difficult to identify until lately in the absence of well-powered family designs^10,12,17^.

The disproportionate study of individuals of EUR ancestries biases the genomic findings and limits their transferability to individuals of different genetic ancestries^26,28–32^. For example, the prediction accuracy of polygenic scores (PGSs) originating from EUR-based GWASs decreases proportionally along the genetic ancestry continuum, as the genetic distance between the training EUR samples and the target samples increases^28,33–35^. Similarly, rare genetic variants associated with disorders or predicted to be pathogenic may not be detected in individuals with non-EUR ancestries due to population specificity or differences in allele frequency and effect size across populations^35–37^. Potential rare variants contributing to disorders in other populations, but absent in EUR ancestries, are missed. In parallel, true benign variants may get misclassified as pathogenic in EUR populations while commonly observed in other ancestries^38^, further hindering interpretation of rare variation, even in the well-studied EUR individuals^39,40^. As a consequence, implementation of genetically informed patient stratification and diagnostics cannot be executed accurately and most importantly equitably, creating significant gaps in precision medicine. Patients with a significant proportion of African (AFR) genetic ancestry, such as African Americans (AfrAms) in the United States, are particularly impacted, as they are genetically distant from the EUR genetic ancestries on the genetic ancestry continuum^33^ and are severely under-represented in neurodevelopmental and neuropsychiatric disorders’ research. Moreover, genetic ancestry varies individual-to-individual within AfrAms due to the admixture process^41,42^, which raises the need for ancestry consideration in studies of this population.

Despite global efforts to correct population imbalance in genomic research^26,39,43–45^, as of September 2025, only 3.87% of participants in the GWAS Catalog^46^ are of AFR ancestry, with 3.71% of them represented by AfrAms and Afro-Caribbeans and 0.16% consisting of continental Africans^27^. In addition to the important mission of ensuring that all patients benefit from advances in genetics and medicine, including non-EUR heterogeneous populations, including admixed ones, in genomic research is advantageous for scientific knowledge and discovery. Indeed, the genomes of individuals from Africa show the highest numbers of variant sites, the lowest degrees of linkage disequilibrium (LD), and the shortest LD block sizes^36^, features that can substantially improve fine mapping of causal variants and power to detect disease variant association^47–50^. Populations from Africa also carry the highest percentages of variants private to the single continent^36^, which facilitates the study of population specific or enriched genetic variation^48^ and can improve understanding of genetic causes of disease, revealing shared and unique genetic effects^47^.

For these reasons, in 2008 we developed the GENAA (Genetics of Neurodevelopment in African Americans) Consortium via an Autism Center of Excellence (ACE) Network to advance this critical research direction and specifically recruit and engage AfrAm families in a nation-wide genetic study of ASD^2,3^. Here, we leverage this resource and present the first comprehensive genetic analysis of AfrAm autistic individuals and their families at the level of both genome-wide (global) and locus-specific (local) genetic ancestry. We performed genotyping and WES on samples from these AfrAm individuals, for whom we confirmed AFR genetically inferred ancestries (GIAs), and compared findings to those from predominantly EUR ASD cohorts. We observe consistent effect sizes of common single nucleotide polymorphisms (SNPs) despite their lack of genome-wide significant association with ASD. We demonstrate the poor performance of existing ASD PGS in AFR GIA individuals, even after consideration of global or local genetic ancestry. We reproduce rare copy number variant (CNV) classes of known high impact as associated with ASD and observe known ASD-associated genes among those impacted by rare variation. We also implicate new candidate genes affected by variants identified in AFR GIA carriers and reveal ASD association for *de novo* missense variants of predicted lower impact. These data and subsequent analyses from GENAA represent a unique resource for gene discovery and a concrete step towards more equitable health, supporting the continued development of larger and more heterogeneous cohorts for genetic research in ASD, and neurodevelopmental and neuropsychiatric disorders more broadly.

## Results

### Genetic ancestry in the GENAA cohort

We recruited individuals with known or suspected ASD and their family members self-identifying as Black or AfrAm in four areas across the country, Bronx, NY, Atlanta, GA, St. Louis, MO, and Los Angeles, CA^2,3^ (Methods). ASD participants were comprehensively evaluated by experienced clinicians from major autism centers, using standard diagnostic tools^2,3^ (Methods). We also obtained DNA samples from selected individuals of the Autism Genetic Resource Exchange (AGRE) cohort (Methods). We generated genotyping and WES data from 2,179 and 1,324 participants, respectively, from GENAA and AGRE to analyze common and rare variants for ASD in AfrAm individuals for the first time (**Fig. 1A**; Supplementary Table 1,2). After quality control (QC) (Methods), genotyping data was available for the analysis of 2,131 individuals (GENAA: n = 1,071; AGRE: n = 1,060), whereas WES data included 1,297 individuals (GENAA: n = 1,118; AGRE: n = 179) (Supplementary Table 1,2).

**Fig. 1.**
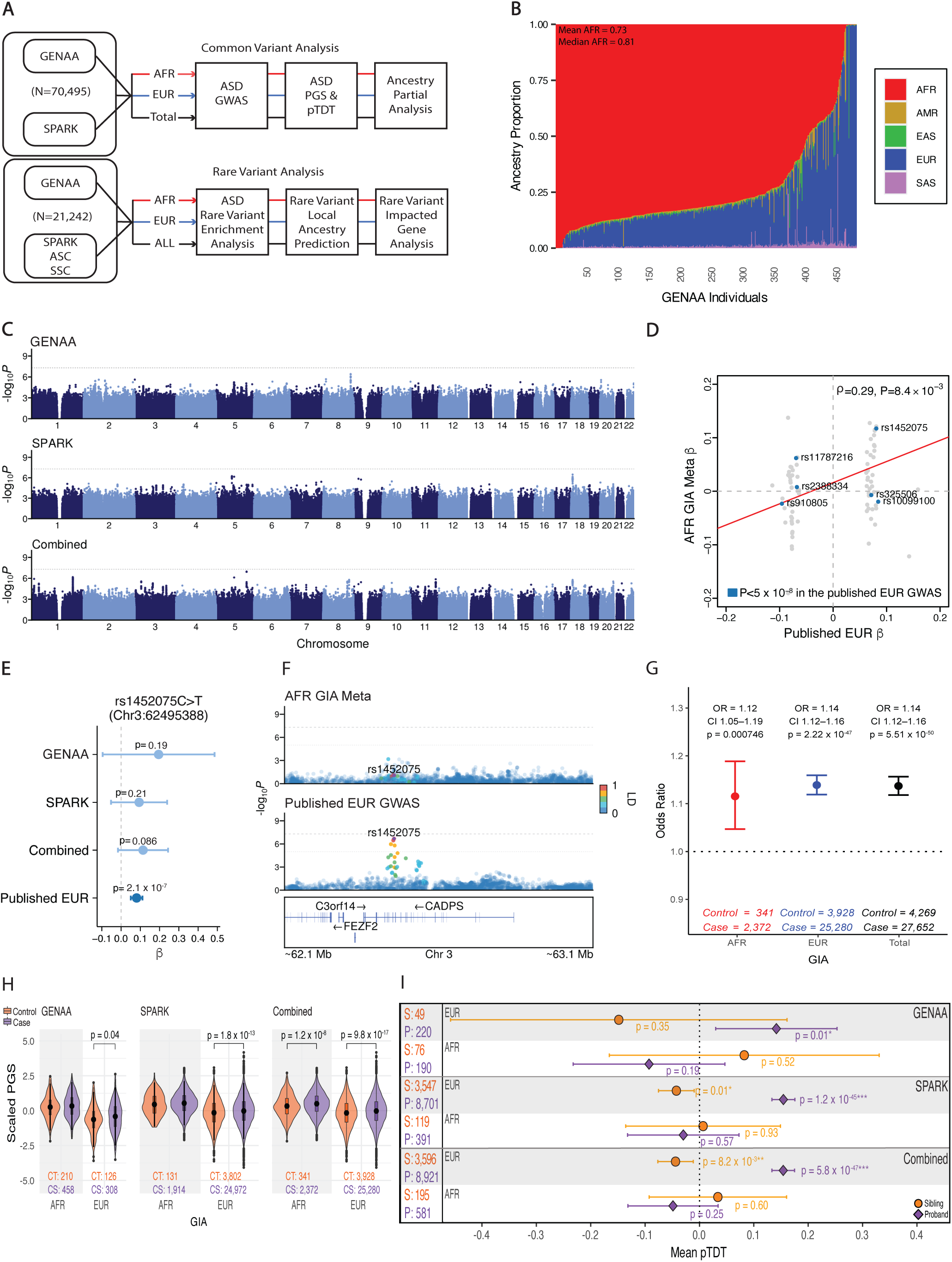
Comparison of AFR GIA based and EUR-based ASD GWASs and performance of EUR-based ASD PGS in AFR GIA samples from the GENAA and SPARK cohorts. (A) Study overview. GENAA genotyping data were combined with the SPARK cohort genotypes for the analysis of common variation in ASD through GWAS, PGS and pTDT, and ancestry partial PGS. Rare variant sets identified from the GENAA WES data were combined with those aggregated from the ASC, SPARK, and SSC cohorts for the investigation of ASD rare variation through rare variant association testing, local ancestry prediction, and impacted gene analysis. Most common variant analyses were performed on three groups of individuals, the AFR GIA and EUR GIA groups (Methods), and the Total group combining them. Most rare variant analyses were performed on family offspring divided in three groups, the AFR GIA and EUR GIA groups (Methods), and the ALL group including individuals with any GIA label. (B) Global genetic ancestry proportions estimated for the GENAA genotyped autistic individuals (excluding AGRE) using the five labeled reference populations from the 1000 Genomes Project. Individuals were ordered on the x axis by decreasing AFR global genetic ancestry proportion. Mean and median AFR global genetic ancestry proportions are reported. (C) Manhattan plots for ASD GWASs on AFR GIA unrelated individuals from GENAA, SPARK, and the two cohorts combined for meta-analysis. The dotted gray horizontal line in each plot indicates the genome-wide significance threshold (*P* = 5 x 10^-8^) for SNP association with ASD diagnosis. (D) Spearman correlation of β coefficients of selected SNPs (Methods) from the EUR-based ASD GWAS^9^, reported on the x axis, and the AFR GIA based GENAA and SPARK ASD GWAS meta-analysis, shown on the y axis. Lead SNPs (R^2^ < 0.1, *P* < 10^-5^) identified in the EUR-based ASD GWAS^9^ were selected for the calculation of the correlation coefficient (Methods). Six of the twelve genome-wide significant SNPs identified in the EUR-based ASD GWAS^9^ are labeled and highlighted in blue (Methods). SNP rs325506^9^ was among the lead SNPs included in the correlation analysis. The other five SNPs^9^ (R^2^ > 0.1, *P* < 10^-5^) were included just for display (Methods). (E) β coefficients and corresponding 95% CIs for chromosome 3 lead SNP rs1452075, previously identified as genome-wide significant in an ASD-Educational attainment combined GWAS analysis^9^, obtained in four different ASD GWASs: the AFR GIA based GENAA, SPARK, and combined meta-analysis GWASs, and the published EUR-based GWAS^9^. P-values from the ASD-only GWASs are shown. (F) Regional association plots around rs1452075 (purple diamond) from the AFR GIA based GENAA and SPARK ASD GWAS meta-analysis (top) and from the EUR-based ASD GWAS^9^ (bottom). SNPs in the shown region are colored by degree of LD to rs1452075 (R^2^), based on the 1000 Genomes Project corresponding reference population (AFR for top plot, EUR for bottom plot) data. Genome-wide (*P* = 5 x 10^-8^) and suggestive (*P* = 10^-5^) significance thresholds are shown. Collapsed gene models for protein-coding genes from GENCODE v42 within the region are displayed. (G) ASD diagnosis ORs and corresponding 95% CIs, and p-values from logistic regression for ASD PGS in three family offspring groups, AFR GIA, EUR GIA, and Total, from the combined GENAA and SPARK cohorts (Methods). Group sample sizes are reported. (H) Distributions of root-mean-square-scaled ASD PGS in autistic (purple) and nonautistic (orange) offspring from GENAA and SPARK by GIA and cohort (Methods). Distributions from the two groups were compared via t-tests. Significant p-values are reported. (I) Mean pTDT deviation values and corresponding 95% CIs in autistic (purple) and nonautistic (orange) offspring from GENAA and SPARK by GIA and cohort (Methods). Significant p-values are denoted with asterisks. Values underlying Fig. 1 are reported in Supplementary Table 5-7,19.

We obtained each individual GIA (Methods) based on their genetic similarity to five reference populations from the 1000 Genomes Project (AFR; AMR: Admixed American; EAS: East Asian; EUR; SAS: South Asian; **Fig. 1B** and **Supplementary Fig. 1A,B,C**). As expected, most (84.4%, n = 407) of the genotyped GENAA-only autistic individuals (n = 482; Supplementary Table 1,2) were confirmed as of AFR GIA (**Supplementary Fig. 1A**), as described in the Methods^51^, with a median AFR global genetic ancestry proportion of 0.81, and a range of 0 to 1, consistent with demographic admixture of AfrAm individuals^41,42,51–53^ and the expected continuum of genetic ancestry (**Fig. 1B**). This trend held true for nonautistic siblings (n = 198; 92.4% of AFR GIA, n = 183; Supplementary Table 1,2) and parents (n = 391; 83.1% of AFR GIA, n = 325; Supplementary Table 1,2) (**Supplementary Fig. 1B**). The entire GENAA-only genotyping cohort, including parents and offspring (n = 1,071), showed a median AFR global genetic ancestry proportion of 0.82 (range: 0, 1) (**Supplementary Fig. 1C**). We combined data from the carefully ascertained GENAA and AGRE cohorts with additional data from a large published cohort, Simons Foundation Powering Autism Research for Knowledge (SPARK)^12^, to increase power and conduct a more comprehensive analysis of the genetic architecture of ASD in this population. We derived GIA labels for SPARK individuals with available genotypes (n = 68,316) (**Supplementary Fig. 1D**; Methods; Supplementary Table 3,4). As expected, the majority of the SPARK cohort (77.5%; n = 52,913) was of EUR GIA (**Supplementary Fig. 1D**). We also identified 3,766 (5.5%) SPARK individuals with AFR GIA that we included to boost statistical power for AFR GIA based common variant analyses (Supplementary Table 3,4), which yielded a total of 2,372 AFR GIA autistic individuals and 341 AFR GIA nonautistic siblings across cohorts (**Fig. 1A**; Supplementary Table 3). We tested common inherited variation and PGS performance, and subsequently analyzed rare copy number variants (CNVs) and *de novo* single nucleotide variants and small indels (SNVs/Indels), to define the genetic architecture of ASD in the GENAA cohort, and in AfrAm individuals from ASD cohorts more broadly (**Fig. 1A**).

### Cross ancestry comparison of common variant genetic effects on ASD

The first common genetic variants robustly associated with ASD were reported in an EUR-based GWAS^9^. To compare with these published data in an EUR cohort^9^, we performed an ASD GWAS with PLINK^54^ using unrelated AFR GIA individuals from GENAA and SPARK (**Fig. 1A**; Methods). As expected (**Supplementary Fig. 1E**), based on the limited power observed in even larger EUR ASD cohorts, we did not find any genome-wide significant (*P* ≤ 5.0 x 10^-8^) associations in the GENAA and SPARK individuals separately, or in meta-analysis (**Fig. 1C**; Supplementary Table 5-7). In addition, we did not observe concordant common variant effect sizes between the GENAA and SPARK analyses when testing the correlation of β coefficients of the top p-value-ranked independent SNPs in common (R^2^ < 0.1 for top one million SNPs, then selection of top 100 SNPs; Methods) between the two datasets (**Supplementary Fig. 1F**; Spearman *ρ* = 0.08, *P* = 0.41).

We used various analytical strategies to increase sample size for the AFR GIA based ASD GWAS and improve its statistical power (**Supplementary Fig. 2**; Methods). First, we used SAIGE^55^, a generalized mixed model that accounts for sample relatedness, to perform ASD GWAS on both unrelated and related AFR GIA individuals from GENAA and SPARK (**Supplementary Fig. 2A**; Supplementary Table 8-10). Second, given the well-documented higher power of quantitative trait GWAS compared to categorical binary GWAS^56–58^, we performed GWAS for the quantitative autistic trait measure Social Communication Questionnaire (SCQ)^59^ raw total score on unrelated and related AFR GIA individuals from GENAA (Supplementary Table 11) and SPARK using GCTA^60^ (**Supplementary Fig. 2B**; Supplementary Table 12-14). Third, we included individuals from the large ABCD^61,62^ cohort (n = 1,953; Methods) with available Social Responsiveness Scale (SRS)^63^ raw total scores, also obtained for the GENAA cohort (Supplementary Table 11), and performed GWAS for this quantitative autistic trait measure on unrelated and related AFR GIA individuals from GENAA and ABCD (**Supplementary Fig. 2C**; Supplementary Table 15-17). We tested the different cohort individuals both separately and in combination and, still, in most analyses except from the SRS GWAS meta-analysis (**Supplementary Fig. 2C**; Supplementary Table 17) and the SPARK SCQ GWAS (**Supplementary Fig. 2B**; Supplementary Table 13), perhaps as expected based on sample size, we did not identify any genome-wide significant association (**Supplementary Fig. 2**).

Next, we contrasted results from our main AFR GIA based ASD GWAS meta-analysis of GENAA and SPARK (**Fig. 1C**) with findings from the EUR-based published ASD GWAS^9^. We compared the effect sizes of defined lead SNPs (*P* < 10^-5^; R^2^ < 0.1; Methods) from the EUR cohort with the AFR GIA based ASD GWAS meta-analysis and found a significant positive correlation (*ρ* = 0.29, *P* = 8.4 x 10^-3^) (**Fig. 1D**). We observed a consistent result (*ρ* = 0.33, *P* = 2.0 x 10^-3^) when comparing effect sizes from the EUR-based study and the AFR GIA based ASD GWAS meta-analysis performed with SAIGE (**Supplementary Fig. 2A**). These data showed moderate sharing of GWAS signal between published EUR cohorts and our GWAS in AFR GIA participants with ASD.

We then specifically examined the genome-wide significant loci from the EUR-based study, with *P* < 10^-5^ in the ASD-only GWAS summary statistics file^9^ (Methods), within the AFR GIA based ASD GWAS in GENAA and SPARK. Among the testable SNPs, we found consistent effect sizes across analyses for index SNP rs1452075 (*P* = 2.1 x 10^-7^)^9^, located on chromosome 3 near the gene *CADPS* (**Fig. 1E**), suggesting its genetic effects for ASD were not discordant across ancestries (**Fig. 1E**; Cochran’s Q test for heterogeneity on Combined and Published EUR; *P* = 0.62; *I*^2^ = 0%; τ^2^ = 0, standard error [SE] = 0.003). We reasoned that the partial GWAS signal replication could be due to shared contributing variants across AFR and EUR chromosomal segments in the two populations^64^, or the effect of EUR admixture in AfrAms^41,42,51–53^. We observed similar LD patterns at the ASD-associated locus (SNP rs1452075) in the AFR and EUR genetic ancestry reference populations from the 1000 Genomes Project (**Fig. 1F**). Then, we performed genetic ancestry deconvolution through RFMix^65,66^ on GENAA and SPARK individuals of AFR, EUR, or admixed AFR-EUR inferred ancestries to identify portions of their genomes of either AFR or EUR origin (**Fig. 1A**; Methods). We used these local ancestry predictions to further investigate the preserved ASD-associated locus (Methods; Supplementary Table 18). We observed both heterozygous and homozygous effect alleles for SNP rs1452075 on an AFR background in autistic individuals and total individuals from GENAA and SPARK included in the AFR GIA based ASD GWASs (Methods; Supplementary Table 18). We then evaluated if in these individuals the SNP resided within a region with AFR/EUR, AFR/AFR, or EUR/EUR diploid local ancestry and tested potential significant deviations from the expected diploid local ancestry frequencies, based on the individuals’ proportions of AFR global ancestry (Methods; Supplementary Table 18). In SPARK, we found that the observed counts for individuals with AFR/EUR, AFR/AFR, or EUR/EUR local ancestry at the locus significantly deviated from the expected values (Chi-squared test; total: *P* = 0.0015; autistic: *P* = 0.02; Supplementary Table 18). In particular, we observed significantly fewer EUR/EUR individuals and slightly more AFR/EUR individuals than expected at this locus. These results indicate that EUR/EUR local ancestry at SNP rs1452075 in AFR GIA individuals tends to be less frequent and suggest that this region is of likely AFR origin and preserved in EUR genomes, which explains the homogeneous effect sizes of the ASD-associated SNP across ancestries.

### Reduced performance of EUR-based ASD PGS in AFR GIA admixed individuals

Having shown partial correlation in individual genetic effects across ancestries, we next analyzed PGS prediction. We evaluated the performance of the most accurate current EUR-derived ASD PGS in the AFR GIA group (**Fig. 1A**) using the SNP association summary statistics from the published ASD GWAS^9,67,68^ in EUR individuals to calculate ASD PGSs in GENAA and SPARK (Methods). As expected, we confirmed that the ASD PGS showed a significant, albeit modest, association with ASD diagnosis (Methods; logistic regression; odds ratio [OR] = 1.14, 95% confidence interval [CI]: 1.12-1.16, *P* = 2.2 x 10^-47^; Supplementary Table 19) in EUR GIA offspring from the combined GENAA and SPARK cohorts (**Fig. 1G**). Although we expected a decrease in the predictive power of the EUR PGS in AFR GIA individuals based on published studies^28,35^, we found a similar association of PGS with ASD in AFR GIA offspring from the combined cohorts (OR = 1.12, 95% CI: 1.05-1.19, *P* = 7.5 x 10^-4^; Supplementary Table 19; **Fig. 1G**); the same observation held true when including both AFR GIA and EUR GIA offspring in the analysis (**Fig. 1G**; OR = 1.14, 95% CI: 1.12-1.16, *P* = 5.5 x 10^-50^; Supplementary Table 19). We also compared ASD PGS distributions between autistic and nonautistic offspring by GIA and cohort (t-test; **Fig. 1H**; Supplementary Table 19). As expected, we found higher ASD PGSs in EUR GIA probands from GENAA (*P* = 0.04), SPARK (*P* = 1.86 x 10^-13^), and the two cohorts combined (*P* = 9.84 x 10^-17^) (**Fig. 1H**; Supplementary Table 19). We observed increased ASD PGS in AFR GIA probands only in the combined cohorts (*P* = 1.2 x 10^-8^), but not separately in either GENAA (*P* = 0.22) or SPARK (*P* = 0.22) (**Fig. 1H**; Supplementary Table 19), consistent with the increased power.

We further investigated ASD PGS transferability to AfrAms by polygenic transmission disequilibrium testing (pTDT)^8^, which evaluates the over transmission of common effect alleles from parents to their offspring, thus controlling for genetic background^8^. We tested pTDT deviation for ASD PGS in both autistic and nonautistic offspring with genotypes available from both parents by GIA and cohort (**Fig. 1I**; Methods). As expected, we found that EUR GIA probands significantly over inherited ASD PGS from their parents (**Fig. 1I**; Supplementary Table 19). We also observed significant under-transmission of ASD PGS in EUR GIA nonautistic siblings in the combined cohorts and in SPARK separately (**Fig. 1I**; Supplementary Table 19). In contrast, in the AFR GIA groups, we observed no significant polygenic transmission disequilibrium for the EUR-based ASD PGS (**Fig. 1I**; Supplementary Table 19) despite its previously observed positive association with ASD (**Fig. 1G**), likely due to reduced power and the potential effect of admixture and varying individual AFR global genetic ancestry proportions in the tested offspring.

We next used the local ancestry predictions from RFMix^65,66^ on GENAA and SPARK individuals of AFR, EUR, or admixed AFR-EUR inferred ancestries (Methods) to compute ancestry partial EUR-based ASD PGSs, PGSs applied only to the EUR or AFR segments of the genome (**Fig. 1A** and **Fig. 2A**; Methods). This allowed the comparison of PGS performance on both backgrounds at the locus level. We evaluated the predictive power of ancestry partial and total ASD PGS for ASD diagnosis through logistic regression (**Fig. 2B**). Similar to the total PGS, the EUR ancestry partial PGS was not a predictor of ASD diagnosis in GENAA (AFR: OR = 0.97, *P* = 0.41; EUR: OR = 1.07, *P* = 0.16; Sum: OR = 1, *P* = 0.92; Supplementary Table 20), likely because of the individual high overall proportion of AFR global genetic ancestry (**Fig. 2B**). In SPARK, which is mostly of EUR GIA, and in the two cohorts combined, we found that both the EUR ancestry partial and total ASD PGSs predicted ASD diagnosis with comparable performance (SPARK - EUR: OR = 1.07, *P* = 3.3 x 10^-12^; Sum: OR = 1.06, *P* = 2.5 x 10^-10^; Combined - EUR: OR = 1.07, *P* = 1.2 x 10^-12^; Sum: OR = 1.06, *P* = 1.1 x 10^-9^; Supplementary Table 20). In contrast, the ASD PGS calculated on the AFR ancestry partial background (the AFR portion of the genome) was not predictive (SPARK - AFR: OR = 0.97, *P* = 0.24; Combined - AFR: OR = 0.97, *P* = 0.20; Supplementary Table 20) (**Fig. 2B**). This analysis suggested that known common variation for ASD identified in primarily EUR populations was not well captured in regions of the genome of AFR genetic ancestry, consistent with findings in other disorders^28,50^.

**Fig. 2.**
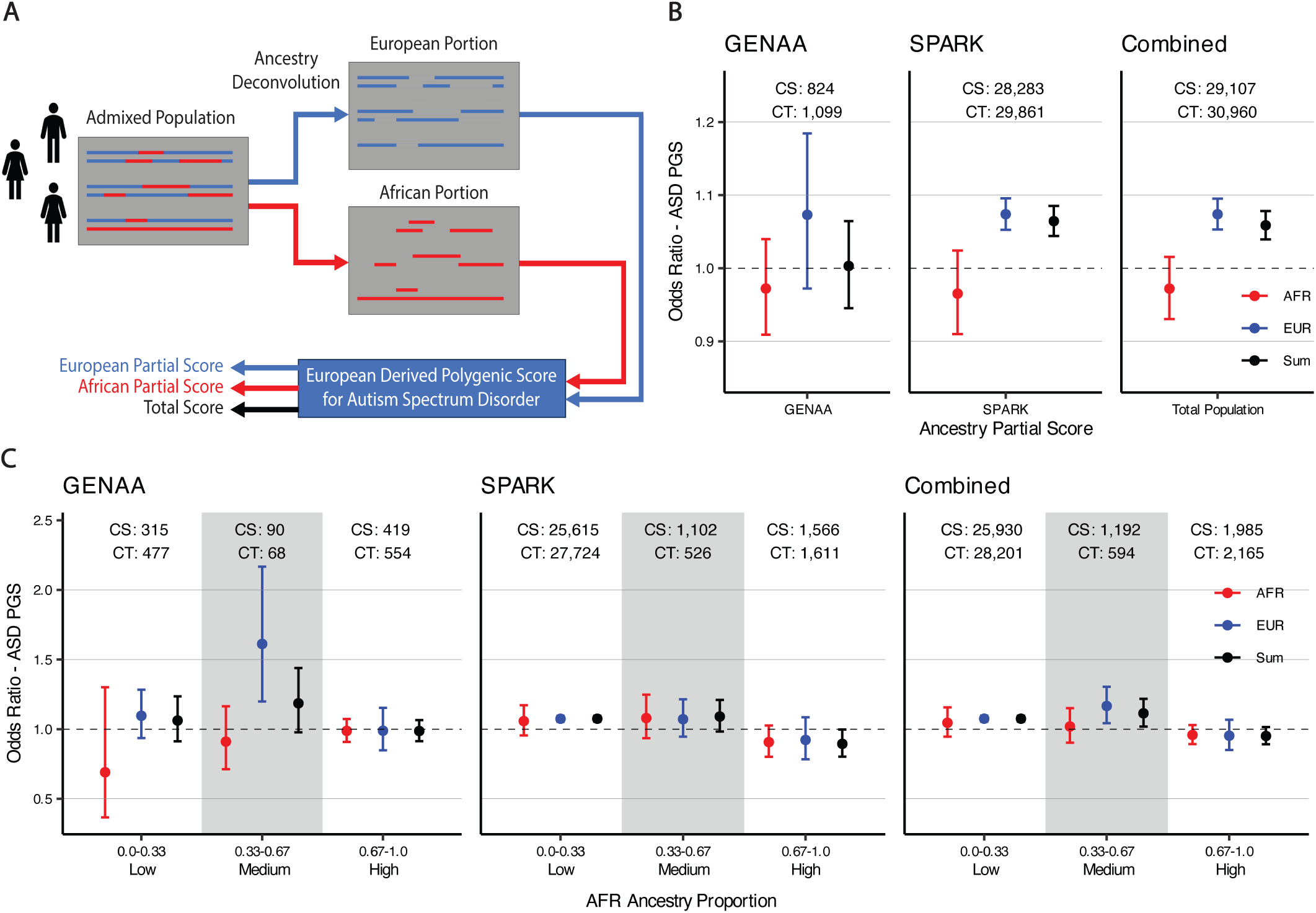
Decreased performance of EUR-based ASD PGS in AFR portions of the genomes of AFR GIA admixed individuals demonstrated through ancestry partial ASD PGSs. (A) Genetic ancestry deconvolution for the computation of ancestry partial ASD PGSs in GENAA and SPARK. The genomes of admixed individuals were compared to both an AFR and EUR reference population to identify which parts came from each ancestral population. An EUR GWAS^9^ based PGS for ASD was then computed for both the AFR and EUR portions of each genome, resulting in both an AFR partial and EUR partial PGS for ASD for every individual. The two partial scores could then be summed to get a per-individual total PGS. (B) ASD diagnosis ORs and corresponding 95% CIs for each ancestry partial ASD PGS (AFR, EUR, and Sum) are shown for individuals, parents and offspring, with AFR, EUR, and admixed AFR-EUR genetic ancestry from GENAA, SPARK, and the two cohorts combined (Methods). Sample sizes for autistic and nonautistic individuals underlying each logistic regression model are reported. (C) ASD diagnosis ORs and corresponding 95% CIs for each ancestry partial ASD PGS (AFR, EUR, and Sum) are shown for individuals of AFR, EUR, or admixed AFR-EUR inferred ancestries from GENAA, SPARK, and the two cohorts combined, grouped into three bins (low, medium, and high) based on their AFR global genetic ancestry proportions (Methods). Sample sizes for autistic and nonautistic individuals underlying each logistic regression model are reported. Values underlying Fig. 2 are reported in Supplementary Table 20.

Given that most individuals in SPARK were primarily EUR, we repeated the analysis by stratifying individuals from each cohort by their proportion of AFR global genetic ancestry (**Fig. 2C**). We observed reduced OR for ASD for both EUR ancestry partial ASD PGS and total ASD PGS in individuals with high proportion of AFR global genetic ancestry compared to those with low proportion in SPARK alone (low AFR ancestry proportion - EUR: OR = 1.07, *P* = 3.1 x 10^-12^; Sum: OR = 1.07, *P* = 1.3 x 10^-12^; high AFR ancestry proportion - EUR: OR = 0.92, *P* = 0.33; Sum: OR = 0.89, *P* = 0.04; Supplementary Table 20) or in the combined cohorts (low AFR ancestry proportion - EUR: OR = 1.08, *P* = 1.4 x 10^-12^; Sum: OR = 1.07, *P* = 7.7 x 10^-13^; high AFR ancestry proportion - EUR: OR = 0.95, *P* = 0.4; Sum: OR = 0.95, *P* = 0.13; Supplementary Table 20) (**Fig. 2C**). Interestingly, we found the EUR ancestry partial ASD PGS as significantly predictive of ASD diagnosis in the GENAA individuals with medium proportion of AFR global genetic ancestry (OR = 1.61, *P* = 0.002; Supplementary Table 20) (**Fig. 2C**). While this group sample size was small, we hypothesized that the signal could be driven by offspring from mixed ancestry couple families. Indeed, we confirmed significant strong enrichment for families of offspring with medium proportion of AFR global genetic ancestry that include one AFR GIA and one EUR GIA parents within the GENAA two-parent families tested (Methods; Fisher’s exact test; *P* = 1.1 x 10^-21^; Supplementary Table 20). Altogether, these findings indicate that the EUR ancestry partial ASD PGS becomes less predictive of ASD diagnosis as a larger portion of the genome is of AFR origin and fewer known ASD common variation can be detected (**Fig. 2C**). In contrast, the AFR ancestry partial ASD PGS showed no association with ASD independently of proportion of AFR origin genome (**Fig. 2C**; Supplementary Table 20), further confirming the reduced transferability of EUR-based findings to genomes of different ancestral origins.

### Consistent excess burden of high-impact CNVs in AFR GIA autistic offspring

We used data from WES to extend our investigation of ASD genetic factors in AfrAms to rare variation (**Fig. 1A**; Supplementary Table 1,2). To maximize statistical power, we combined the rare CNVs and the *de novo* SNVs/Indels identified in the GENAA probands and nonautistic siblings (Methods) with those found in fully phase-able autistic and nonautistic offspring from three large ASD cohorts, the Autism Sequencing Consortium (ASC), SPARK, and the Simons Simplex Collection (SSC)^13^ (**Fig. 1A**; Supplementary Table 21). We divided these individuals into three groups for rare variant analysis: the AFR and the EUR GIA groups, defined by genetic similarity to labeled reference populations using WES data (Methods), and the group including all offspring available for analysis, independent of their genetic ancestry labels (ALL) (**Fig. 1A**; Supplementary Table 21). GENAA comprised 513 autistic probands and 152 nonautistic siblings who passed QC and were confirmed as of AFR GIA (Supplementary Table 1,2). In contrast, of the 20,526 individuals available from the other three ASD cohorts, 15,050 (73.3%) were of EUR GIA, whereas only 700 (3.4%) were of AFR GIA and could be combined with the GENAA cohort for analysis (Supplementary Table 21). Therefore, through the GENAA WES dataset generated in this study, we nearly doubled the current AFR GIA sample size for rare variant analysis.

We evaluated the association of specific classes of rare variants with ASD in each group through variant burden analysis (**Fig. 1A**; Supplementary Table 21). We found consistent excess burden of rare deletions impacting highly constrained genes (with corresponding loss-of-function observed/expected upper bound fraction [LOEUF]^69^ score in the 1^st^ decile) and rare duplications larger than 1 Mb in the autistic offspring compared to the nonautistic offspring across all the three tested groups, AFR, EUR, and ALL (binomial test; **Fig. 3A,B**; Supplementary Table 22). However, autistic probands in the EUR and ALL groups carried significantly more rare duplications in highly constrained genes (1^st^ LOEUF score decile) and rare deletions larger than 1 Mb than the nonautistic siblings (binomial test; **Supplementary Fig. 3A,B**; Supplementary Table 23). Similarly, we found that *de novo* protein-truncating variants (PTVs) in highly constrained genes (1^st^ and 2^nd^ LOEUF score deciles) and high-impact *de novo* missense variants (Missense badness, PolyPhen-2, and Constraint [MPC]^70^ score ≥ 2; Methods) were significantly enriched in autistic offspring compared to nonautistic offspring in the EUR and ALL groups, but not in the AFR GIA group, likely due to small variant counts and the resulting reduced statistical power in the latter (binomial test; **Supplementary Fig. 3C,D**; Supplementary Table 23). These findings confirm that some well-known ASD association signals, first identified in EUR-based analyses, such as those for high-impact rare CNVs^13^, can be replicated in AfrAms. Additionally, in the combined ALL group, we found significant excess burden of *de novo* PTVs in genes under moderate constraint (3^rd^ LOEUF score decile) and of *de novo* missense variants of lower impact (with MPC score < 1) (binomial test; **Supplementary Fig. 3C,D**; Supplementary Table 23; Methods). We did not observe this in the EUR GIA group, consistent with the reduced accuracy of standard functional severity metrics and their thresholds (which are based on EUR populations) in non-EUR, more genetically heterogeneous cohorts^38,40^.

**Fig. 3.**
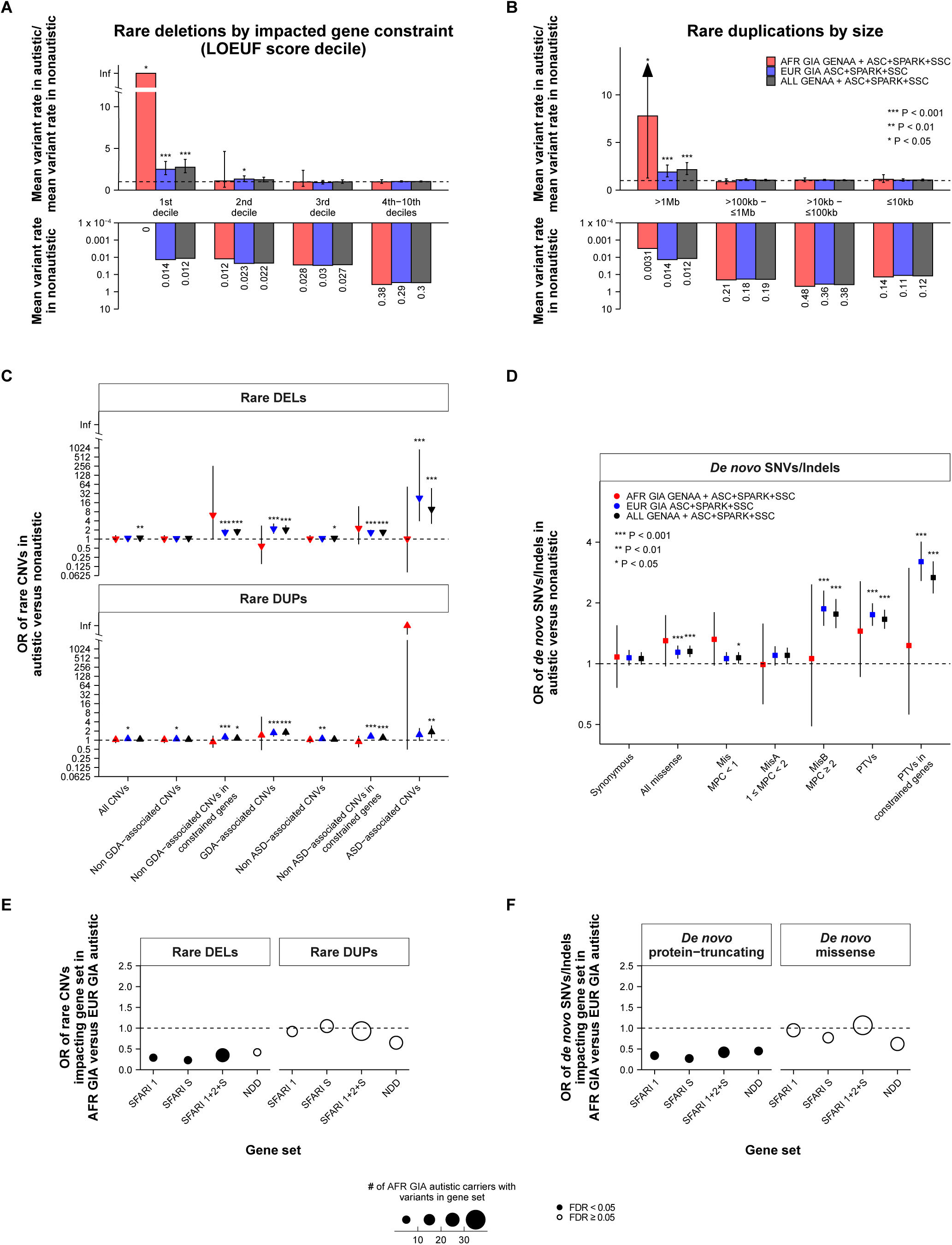
Analysis of rare CNVs and *de novo* SNVs/Indels and their impacted genes in AFR GIA family offspring from the GENAA, ASC, SPARK, and SSC cohorts reveals consistent ASD association signals and potential for discovery. (A-B) Bar plots showing the autistic by nonautistic mean variant rate ratios (top) and the nonautistic mean variant rates (bottom) of rare deletions by LOEUF score decile of the impacted gene (A) and of rare duplications by their size category (B) in three offspring groups based on GIA: AFR (in red), EUR (in blue), and from any GIA (ALL, in black). ASD excess rare variant burden was tested through two-sided one-sample proportion tests via binomial tests on the observed variant counts and proportions of autistic offspring within each tested group. Asterisks show different nominal significance ranges. Error bars represent the 95% CIs for the variant rate ratios, computed through binomial tests. The lower bar plot reversed y axes were log_10_ transformed for visualization. (C-D) Enrichment of autistic carriers of different classes (Methods) of rare CNVs (deletions or DELs and duplications or DUPs) (C) and *de novo* SNVs/Indels (D) within each tested offspring group, AFR, EUR, and ALL. ORs from two-sided Fisher’s exact tests are shown on the y axes, log_2_ transformed for visualization, along with their 95% CIs. Asterisks show different nominal significance ranges. (E-F) Enrichment of AFR autistic carriers of rare CNVs (E) and *de novo* SNVs/Indels (F) impacting genes from specific gene sets, relevant to ASD (Methods), within AFR and EUR autistic carriers. ORs from two-sided Fisher’s exact tests are shown on the y axes. Solid circles represent significant ORs with FDR < 0.05 (per variant set FDR values). Circle size reflects the range for number of AFR autistic carriers of rare CNVs or *de novo* SNVs/Indels impacting each gene set tested. Group sample sizes and test results for analyses underlying Fig. 3 are reported in Supplementary Table 22.

Since matched genotyping and WES data were available for some individuals of the GENAA cohort (Supplementary Table 1,2), we tested if specific rare variant sets of interest from GENAA were more likely to be on a local AFR background than expected. We used local genetic ancestry assignments obtained from genotyping data (Methods) to annotate whether rare variants in GENAA offspring were within AFR or EUR genomic regions, which was limited to those with homozygous local ancestry (**Fig. 1A**; Methods) (**Supplementary Fig. 4**; Supplementary Table 24; Methods). We found no differences in the numbers of variants per Mb between genomic regions with AFR or EUR origin (paired t-test) for both the rare CNVs (**Supplementary Fig. 4A,B**; Supplementary Table 24) and *de novo* SNVs/Indels (**Supplementary Fig. 4C,D**; Supplementary Table 24) for all, autistic, or nonautistic offspring, consistent with expectations. We next focused on a set of most likely rare inherited variants, which we hypothesized would be more likely of AFR origin, by testing CNVs with maximum site frequency across genetic ancestry groups (grpmaxSF) of 1-5% in the Genome Aggregation Database (gnomAD)^39^ (**Supplementary Fig. 4E,F**; Supplementary Table 24; Methods). As hypothesized, we found these rare, but likely inherited CNVs, to be located within regions of AFR homozygous local ancestry compared to those of EUR origin for all GENAA offspring and for probands alone (paired t-test; all: FDR = 0.016; autistic: FDR = 0.033; **Supplementary Fig. 4E,F**; Supplementary Table 24).

### AFR GIA based rare CNV analyses reveal new candidate deletions and genes

We next tested the enrichment of autistic offspring among carriers of specific classes of rare CNVs from each group (AFR, EUR, and ALL) (**Fig. 1A**). We annotated the identified rare CNVs for their overlap with loci known to be associated with ASD and other genomic disorders (GDs) (Supplementary Table 25). This allowed their classification into specifically ASD-associated and broader genomic disorder and/or autism-associated (or GDA-associated) events (Methods). As expected, we observed significant enrichment of ASD-associated and GDA-associated rare deletions and duplications in autistic offspring of the ALL and EUR groups, with the exception of the ASD-associated rare duplications in EUR (Fisher’s exact test; **Fig. 3C**; Supplementary Table 22). The ASD-associated rare deletions were the most ASD-enriched rare CNV class in both groups (ALL: OR = 9.7, *P* = 5.5 x 10^-8^; EUR: OR = 22.6, *P* = 1.3 x 10^-7^; Supplementary Table 22). Consistent with previous investigations and CNV classifications^13^, rare deletions and duplications not corresponding to any ASD-associated or GDA-associated loci, but selected for their impact on at least one constrained gene (LOEUF score < 0.4) also showed significant enrichment in autistic offspring in both the ALL and EUR groups (Fisher’s exact test; **Fig. 3C**; Supplementary Table 22). Intriguingly, we observed a strong, but non-significant trend for non GDA-associated rare deletions impacting at least one constrained gene in AFR GIA probands (Fisher’s exact test; OR = 6.2; *P* = 0.06; **Fig. 3C**; Supplementary Table 22).

We further investigated this potentially novel (non GDA-associated) set of 20 rare deletions impacting at least one constrained gene in AFR GIA carriers through local genetic ancestry and impacted gene analyses (Supplementary Table 26,27; Methods). Inspection of local ancestry did not reveal a significant preference for either AFR or EUR genomic regions in the 12 deletions from GENAA and SPARK for which we were able to assign local ancestry (Methods; paired t-test; **Fig. 4A** and **Supplementary Fig. 5A**; Supplementary Table 28).

**Fig. 4.**
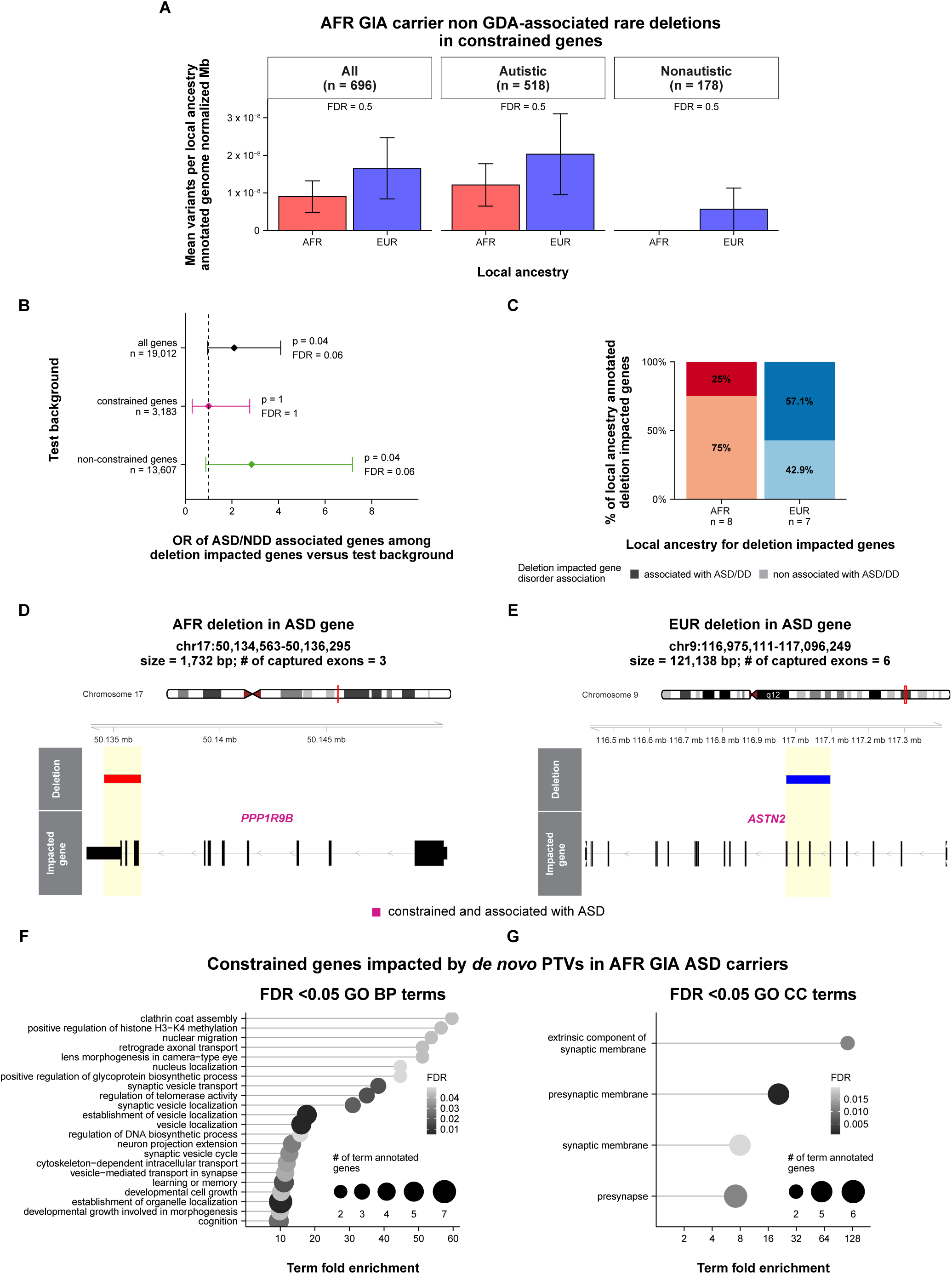
Local genetic ancestry and impacted gene analysis of the potentially novel non GDA-associated rare deletions in constrained genes identified in AFR GIA carriers and GO term enrichment analysis of the constrained genes impacted by *de novo* PTVs in AFR GIA autistic carriers. (A) Bar plots showing the mean numbers of variants per local ancestry annotated genome normalized Mb for genomic regions with AFR (red) or EUR (blue) (both homozygous and heterozygous) local ancestry in three groups of combined GENAA and SPARK AFR GIA offspring with inferred local genetic ancestry (Methods): all, autistic, and nonautistic. Error bars represent the mean ± standard error values. Per variant set FDR values (controlling for the three simultaneous comparisons) from two-sided one-sample paired t-tests are reported. (B) Enrichment of deletion-impacted genes associated with ASD and/or NDD within three different gene sets used as test backgrounds: all the analyzed genes, only the constrained ones, and just the non-constrained ones (Methods; the number of genes in each test background are reported). ORs from two-sided Fisher’s exact tests are shown on the x axis along with their 95% CIs. Both the nominal p-values and FDR values (for the three simultaneous tests) are reported. (C) Stacked bar plots showing the percentages of genes impacted by the GENAA and SPARK AFR GIA carrier deletions with AFR (red) or EUR (blue) local ancestry (Methods) that are associated (darker shade of red/blue) or not associated (lighter shade) with ASD and/or DD. (D-E) Visualization of the genomic regions capturing two deletion events and their corresponding impacted gene for two GENAA AFR GIA autistic carriers (Methods). Both deletions, one with AFR (D) and one with EUR (E) local ancestry, impact constrained genes that have already been associated with ASD (*PPP1R9B*, with a SFARI Gene score of 1; *ASTN2*, with a SFARI Gene score of 2). Deletion genomic coordinates, sizes, and numbers of impacted well-captured exons are shown. (F-G) Lollipop plots showing the fold enrichment (x axis) (Methods) of significant GO biological process (BP) (F) and cellular component (CC) (G) terms (y axis) for the list of constrained genes impacted by *de novo* PTVs in at least one AFR GIA autistic carrier. The dot size corresponds to the number of list genes annotated with each specific term, whereas the dot color reflects the one-sided Fisher’s exact test FDR value (Methods). The x axis in (G) was log_2_ transformed for visualization. Analysis values underlying Fig. 4 are reported in Supplementary Table 28.

Next, to understand why this potentially novel deletion set was enriched in autistic offspring, we verified if the 20 rare deletions impacted genes that had already been associated with ASD, other neurodevelopmental disorders (NDDs), and/or general developmental disorders (DDs) (Methods; Supplementary Table 27). We tested the enrichment of known ASD and NDD associated genes (Methods) among the deletion-impacted genes against three different background gene sets, (1) the entire gene set used for CNV calling, and its two (2) constrained and (3) non constrained gene subsets (Methods) (Fisher’s exact test; **Fig. 4B**; Supplementary Table 28). We observed nominally significant enrichment of known ASD and/or NDD-associated deletion-impacted genes within both all and the non-constrained genes analyzed (Fisher’s exact test; all genes - OR = 2.1, *P* = 0.04; non constrained genes - OR = 2.8, *P* = 0.04; **Fig. 4B**; Supplementary Table 28). This enrichment signal was absent within constrained genes, likely due to the small numbers tested (Fisher’s exact test; OR = 1.0, *P* = 1; **Fig. 4B**; Supplementary Table 28). Indeed, we observed no difference between the proportions of genes associated with ASD, NDD, and/or DD among the constrained and non-constrained genes impacted by the 20 deletions (Pearson’s chi-squared test; *P* = 0.9; **Supplementary Fig. 5B**). Therefore, we confirmed that this deletion class, initially defined by gene constraint, was enriched in AFR GIA probands likely due to its significant impact on known developmental disorder-associated genes as derived from current databases. Interestingly, despite some of the deleted genes being linked to ASD and/or NDD, these deletion loci had not already been confidently associated with ASD or other GDs.

Next, we asked if known disorder genes were more frequently deleted on either AFR or EUR chromosomal backgrounds to highlight any potential bias due to the population imbalance in genetic research; given the insufficient power, we found no significant enrichment for known ASD and DD associated genes impacted by deletions from this set located within EUR genomic segments (**Fig. 4C**; Fisher’s exact test; OR = 3.6; *P* = 0.3; Supplementary Table 28). As expected, we found known ASD genes, such as *PPP1R9B* and *ASTN2*, impacted by deletions in this set, independent of their local ancestry (**Fig. 4D,E**; Supplementary Table 26,27). Similarly, we observed that these deletions impacted possibly new genes, which are constrained, but have not been associated with neurodevelopmental disorders yet, like *SMC2*, *FBLN5*, *DMTN*, *SORCS1*, and *GLYR1*, or are not constrained, but have been linked to recessive severe developmental disorders, such as *TRIP11* and *ROGDI* (**Supplementary Fig. 5C,D**; Supplementary Table 26,27). Importantly, the potentially novel genes represented a large proportion (75.3%) of the constraint annotated genes deleted in these AFR GIA carriers (**Supplementary Fig. 5B**), which suggests that there is opportunity for gene discovery with the analysis of AfrAms and more patients from a variety of backgrounds.

### *De novo* protein-truncating and missense variants in AFR GIA autistic offspring

We tested association of specific classes of *de novo* PTVs and missense variants with ASD. Reproducing results from previous studies^11–13^, we found significant ASD enrichment for all *de novo* missense variants with MPC score annotation, high-impact missense variants (MisB variants, with MPC score ≥ 2), PTVs, and PTVs within constrained genes (LOEUF score < 0.4) in both the ALL and EUR groups (Fisher’s exact test; **Fig. 3D**), with the last variant class showing the strongest signal among the *de novo* SNVs/Indels (ALL: OR = 2.7, *P* = 2.8 x 10^-33^; EUR: OR = 3.2, *P* = 3.6 x 10^-33^; Supplementary Table 22). While not reproducing the expected ASD association of *de novo* PTVs in constrained genes in the AFR GIA group (**Fig. 3D**; **Supplementary Fig. 3C**; Supplementary Table 22,23), we confirmed through gene ontology (GO) term enrichment analyses (Methods; Supplementary Table 28) that the constrained genes impacted by *de novo* PTVs in AFR GIA autistic carriers were involved in biological processes previously shown to be impacted in ASD, such as neurotransmission and chromatin remodeling^10,12,71,72^ (**Fig. 4F**). As expected, these genes encoded presynaptic membrane components (**Fig. 4G**; Supplementary Table 28) and were confirmed to be annotated as ASD genes in the Disease Ontology (DO)^73^ annotation term database (**Supplementary Fig. 6A**; Supplementary Table 29; Methods). These observations indicate that the genes and biological processes impacted in ASD are shared across ancestries, as would be expected based upon shared biology. Interestingly, in agreement with corresponding findings from the rare variant burden analyses (**Supplementary Fig. 3D**; Supplementary Table 23), we found that *de novo* lower-impact missense variants (MPC score < 1) were significantly enriched in probands in the ALL group (OR = 1.1, *P* = 0.04; Fisher’s exact test; **Fig. 3D**; Supplementary Table 22) and showed slightly higher but non-significant trend towards ASD enrichment in the AFR GIA group, which drives this novel ASD association signal (OR = 1.3, *P* = 0.07; Fisher’s exact test; **Fig. 3D**; Supplementary Table 22). As expected, when testing the local ancestry of the GENAA *de novo* missense variant set (**Supplementary Fig. 4G,H**), we found no differences in the numbers of variants per Mb between genomic regions with either AFR or EUR origin (paired t-test; Supplementary Table 24). These observations highlight how the analysis of genetically heterogenous individuals such as AfrAms may also offer new insights for the understanding of rare variation and the potential refinement of functional severity measures and their thresholds.

### Depletion of rare variants in known associated genes in AFR GIA autistic carriers

Next, we verified if the rare variants found in AFR GIA probands impacted known associated genes identified in cohorts of EUR ancestry (**Fig. 1A**). We curated four gene sets, selected for their relevance to ASD, for this analysis: Simons Foundation Autism Research Initiative (SFARI) 1 (high-confidence ASD genes), SFARI S (syndromic ASD genes), SFARI 1+2+S (union of the high-confidence, strong candidate, and syndromic ASD genes), and NDD (neurodevelopmental disorder genes) (Methods). First, we confirmed significant enrichment of autistic offspring among carriers of rare deletions and duplications (**Supplementary Fig. 3E**) and *de novo* PTVs and missense variants (**Supplementary Fig. 3F**) impacting each of the four gene sets consistently in both the ALL and EUR groups (Fisher’s exact test; Supplementary Table 23). The strongest ASD enrichment was for the rare deletions in the NDD gene set and for the *de novo* PTVs in the SFARI 1, NDD, and SFARI S gene sets (Fisher’s exact test; Supplementary Table 23). Only rare duplications impacting SFARI S genes were significantly enriched in probands consistently for all the three analysis groups, AFR, EUR, and ALL, with the strongest OR of 4.6 in the AFR GIA offspring (FDR = 0.03; Fisher’s exact test; Supplementary Table 23). GO term enrichment analyses confirmed that the known ASD and NDD genes impacted by rare variants in AFR GIA autistic carriers were involved in neurotransmission, cognitive processes, and chromatin remodeling (**Supplementary Fig. 6B,C**; Supplementary Table 29), and were linked to ASD, intellectual disability, and rare genetic conditions (**Supplementary Fig. 6D**; Supplementary Table 29). In addition, based on analyses using both the Disease Association Protein-Protein Link Evaluator (DAPPLE)^74^ and STRING^75^ (Methods), these genes formed a significant protein-protein interaction (PPI) network, which further demonstrates their biological connections to known ASD-associated pathways (DAPPLE: direct edge count *P* = 0.003, average indirect connectivity *P* = 0.01; STRING: PPI enrichment *P* = 0.0004).

We then asked if carriers of rare variants impacting the four gene sets were enriched in either AFR GIA or EUR GIA autistic carriers of rare CNVs and *de novo* PTVs and missense variants. Rather than seeing an equal distribution across ancestries, we observed a significant depletion of rare deletions and *de novo* PTVs in known ASD genes (SFARI 1, SFARI S, and SFARI 1+2+S) in AFR GIA probands (Fisher’s exact test; **Fig. 3E,F**; Supplementary Table 22). The same significant pattern was also observed for the *de novo* PTVs in known NDD genes (**Fig. 3F**; Supplementary Table 22). Therefore, loss-of-function mutations in known associated genes were less frequent in AFR GIA probands than EUR GIA probands. We next identified constrained genes impacted by rare deletions and *de novo* PTVs in at least one AFR GIA autistic carrier that were not part of the tested gene sets, and therefore potentially new (n = 37; Methods), and found that they encoded components of the synaptic membrane crucial for neurotransmission (Supplementary Table 29). Overall, these findings suggest that new genes, likely involved in known mechanisms altered in ASD, may be discovered through the analysis of more AfrAms and other participants with AFR and other non-EUR ancestries.

## Discussion

Here we describe the genetic architecture, including common and rare genetic factors, in AfrAms with ASD for the first time. We leverage our GENAA Consortium and report the first genetic analysis from this AFR GIA admixed cohort. We showed consistent cross ancestry effect sizes of common variants for ASD, but overall reduced performance of current ASD PGS in AFR GIA individuals; this is largely due to the existing ASD PGS being primarily trained in EUR ancestry populations. Moreover, by deconvolution of local genetic ancestry, we were able to assess genetic contributions at locus-specific ancestry resolution. We identified a depletion in loss-of-function mutations in known associated genes and promising candidate loci for ASD based on the observed strong trend for novel, previously non GDA-associated rare deletions in constrained genes in AFR GIA autistic individuals. We recognize that, compared with EUR cohorts, our cohort size is more modest, but our results strongly emphasize the value of increasing the sample size of non-EUR populations, especially AfrAms or other AFR GIA populations, for gene discovery due to their increased genetic diversity, and for equitable healthcare based on PGS. Through this work, we generated, described, and share an impactful resource, which provides a starting point for the potential of making genetic medicine in ASD, and neurodevelopmental and neuropsychiatric disorders broadly, more equitable.

Through the meta-analysis of AFR GIA individuals in the GENAA and SPARK cohorts, we demonstrated that, despite the expected lack of significant genome-wide associations, the effect sizes of lead SNPs from the published EUR ASD GWAS were positively correlated with those in the AFR GIA based ASD GWAS. This finding suggests that ASD common variation is at least partially shared across genetic ancestry groups. In particular, for one common SNP, rs1452075, previously identified as strongly associated with ASD in EUR cohorts, we confirmed concordant effect sizes in AFR GIA individuals. Moreover, by local ancestry analysis we demonstrated that this locus is likely of AFR origin, and we showed its preserved LD structure between AFR and EUR ancestry genomes, which explains the ASD effect consistency. This highlights the importance of analyzing more genetically heterogeneous individuals to improve knowledge of LD patterns, design of genotyping arrays, performance of genotype imputation, and, ultimately, identification of causative SNPs in individuals from different genetic backgrounds^26,28^.

Unexpectedly, we found some contrasting results on the performance of EUR-based ASD PGS when this was computed for either AFR GIA individuals from the combined GENAA and SPARK cohorts or for chromosomal segments of AFR and EUR origin in individuals with a high proportion of AFR global genetic ancestry from the same cohorts. These results confirm the importance of increased resolution offered by local ancestry prediction in the analysis of admixed populations. Interestingly, the ancestry partial PGS analysis findings were consistent with the pTDT analysis in which we showed, despite the reduced power, that currently known common variants contributing to ASD were not over-transmitted to AFR GIA probands and therefore, could not be reliably linked to ASD in this group defined by GIA. Additionally, through local ancestry deconvolution and in-depth ancestry partial PGS analysis, we found that the EUR-based ASD PGS, beyond performing poorly in AFR genomic regions, showed reduced performance even on EUR segments as their proportion decreased in individuals’ genomes. Altogether, these results confirm the non-transferability of EUR-derived findings and tools based on common variation, such as PGS, in other genetic ancestries. Overall, they stress the need for better powered ASD GWAS in non-EUR cohorts in addition to the work being done in EUR populations^9,48–50^ and highlight the potential of local ancestry deconvolution for the optimization of PGS.

Through the investigation of rare variation and its impacted protein-coding genes in AFR GIA offspring from four combined ASD cohorts, nearly half of whom were recruited in GENAA, we collected evidence supporting two key conclusions. The first is that major effect size variant classes identified in cohorts of EUR ancestry can be replicated in those with AFR GIA, as previously hypothesized^40,76^. Rare variants of large effect size are expected to be at least partially shared and transferable across ancestries, despite differences in allele frequencies and the existence of population-specific variants^36^. Indeed, in our AFR GIA combined cohort we found: (1) consistent excess burden of rare deletions in highly constrained genes and large rare duplications in autistic individuals; (2) that some known ASD, NDD, and DD genes were among those impacted by rare variants and could be observed on both AFR and EUR backgrounds; and (3) that altered biological processes and networks were shared across ancestries, as confirmed by our analysis of *de novo* PTVs in constrained genes.

The second major conclusion is that the investigation of ASD genetic architecture in AfrAms, and for that matter in other genetically heterogeneous cohorts, should be leveraged to facilitate discovery of new ASD-associated genes. We observed a strong near-significant enrichment trend for novel non GDA-associated rare deletions in constrained genes in AFR GIA probands. Intriguingly, among the impacted genes, on either AFR or EUR genomic segments, we noticed several promising new genes based on their constraint or previous association with recessive forms of developmental disorders. In general, these novel rare deletions observed in AFR GIA carriers affected genes that were mostly not already tied to ASD or other developmental disorders. One specific gene, *ROGDI* (rogdi atypical leucine zipper), was deleted on the EUR portion in two AFR GIA autistic siblings from the same family. This gene is involved in brain development and neurogenesis^77^ and implicated in the recessive Kohlschutter-Tonz syndrome, a disorder characterized by seizures, severe global developmental delay, spasticity, and amelogenesis imperfecta^77,78^. The two autistic siblings, who showed severe language delay and ASD symptoms, carried the heterozygous deletion and no other compound heterozygous rare variant at this locus. We also found two specific constrained genes, *SMC2* (structural maintenance of chromosomes 2) and *FBLN5* (fibulin 5), impacted by two deletions on AFR background in the same autistic AFR GIA carrier, who showed moderate ASD symptoms. *SMC2* encodes a molecular component of the condensin complexes that regulate mitotic chromosome condensation and facilitate cell division^79^. Interestingly, other proteins in these complexes have been linked to neurodevelopmental syndromic features, such as microcephaly and mild intellectual disability^80,81^, and even to ASD^12^. *FBLN5* encodes an extracellular matrix protein essential for elastic fiber formation and heterozygous mutations in this gene have been associated with demyelinating Charcot-Marie-Tooth disease type 1H, a slowly progressive peripheral sensorimotor neuropathy with adult onset^82,83^. Moreover, we observed two additional specific constrained genes, *DMTN* (dematin actin binding protein) and *SORCS1* (sortilin related VPS10 domain containing receptor 1), deleted on EUR and AFR background, respectively, in two other AFR GIA autistic carriers. *DMTN* encodes an actin-bundling protein of the erythroid membrane skeleton abundantly expressed in the human brain^84,85^. Intriguingly, despite not being a known ASD-associated gene, *DMTN* has been previously reported as top hub gene of a neuronal co-expression module, representing genes involved in synaptic vesicle transport, that was downregulated in ASD brains^86^. It has been suggested that this module downregulation could be compensated by the ASD brain hyperacetylation of gene linked distal enhancer regions, which were also enriched for ASD common variation^86^. *SORCS1* encodes a neuropeptide receptor that is strongly expressed in the brain^87^ and is involved in synaptic receptor trafficking and synaptic transmission^88^. This gene is localized on chromosome 10 nearby another member of this gene family, *SORCS3* (sortilin related VPS10 domain containing receptor 3), which is a known ASD-associated gene^89^. Overall, the potentially new genes found deleted in autistic carriers of AFR GIA show credible biological connection to already known ASD genes and mechanisms.

Through local ancestry deconvolution we observed that when these new deletions affected known ASD and/or DD genes they tended to occur, even if not significantly, on EUR genomic segments. Consistent with this observed potential bias, despite the caveat of unbalanced carrier group sizes and reduced power in the AFR GIA group, we found that rare deletions and *de novo* PTVs in known ASD and NDD genes were less frequent in AFR GIA autistic carriers compared to those of EUR GIA. This finding suggests that the current understanding of ASD genetic architecture may be excessively tied to the predominant study of EUR ancestry patients^26,28^. In parallel, our data suggests modest enrichment of *de novo* missense variants predicted to be of low impact based on the analysis of primarily EUR genomes^70^ within AFR GIA probands from the four combined ASD cohorts. This is consistent with emerging evidence that more genetically heterogeneous cohorts are needed to obtain accurate measures of variant pathogenicity^37–40,90–92^. Each of these findings emphasizes the need for substantial efforts for further investigation and validation in larger and better-powered sample sizes. Yet, despite the limitations inherent in our study sample size, our findings highlight the potential for discovery through the analysis of AfrAms, as well as other under-studied populations, in neurodevelopmental and neuropsychiatric disorder research and more broadly in biomedicine.

## Acknowledgements

We thank the families participating in GENAA for their contribution to autism genomic research. This work was supported by the National Institute of Mental Health (NIMH; R01MH100027). Data and/or research tools used in the preparation of this manuscript have been deposited to the National Institute of Mental Health Data Archive (NDA). NDA dataset identifiers are listed in the manuscript Data Availability Statement. We thank the individuals from AGRE who participated in this study. We are grateful for the resources from SFARI (for the SPARK and SSC cohorts) and the ASC, which allowed us to expand our investigation of ASD genetic architecture. We thank the UCLA NeuroGenomics Core (UNGC) and the Genomics Core of the Institute of Human Genetics (IHG) at UCSF for their support in genetic data generation. We thank the Terra Support team at the Broad Institute for their technical assistance on the platform. We are grateful to Jessica Way from the Data Sciences Platform at the Broad Institute for the support with WARP editing and testing. We thank David Keller from NIMH Repository & Genomics Resource (NRGR, now Sampled) for the assistance with sample management and acquisition.

## Author Contributions

D.H.G. provided funding for the GENAA recruitment and study. D.H.G., J.N.C., and A.K. designed the GENAA study. S.M., C.K., A.K., N.M., J.N.C., A.M.A., M.F.A., E.T.G., A.N.C., and E.O.A. contributed to recruitment and outreach for GENAA. A.C.G., A.M.A., J.M., A.D.B., S.M., T.H., R.B.W., and T.R.B. performed the GENAA clinical assessments. J.K.L. and Y.Z. contributed to the GENAA clinical data curation and management. J.K.L., H.C., and J.N.H. contributed to genetic data generation from GENAA. M.C. and J.K.L. managed the GENAA genetic data with assistance from S.A.A., R.M.K., and J.E.P.V.. J.K.L., J.N.H., A.O.F.C., and M.E.K. performed processing and QC of the GENAA genotyping data. A.O.F.C., K.H., and V.P. predicted genetically inferred ancestry and local ancestry from the genotyping data under supervision from B.P. and D.E.A.. D.H.G., B.P., D.E.A., R.M.C., L.M.H., and S.J.S. supervised the common variant analyses. M.E.K. and A.O.F.C. performed the GWAS analyses. D.S.G. and A.O.F.C. performed the ASD PGS and pTDT analyses. A.O.F.C. and K.H. performed the ancestry partial ASD PGS analyses. M.C., S.A.A., R.M.K., J.E.P.V., and J.K.L. contributed to processing and QC of the GENAA WES data. S.D. performed *de novo* SNV/Indel calling and QC with supervision from S.J.S.. J.M.F. performed rare CNV calling and QC, with supervision from M.E.T. and assistance from M.C. and J.E.P.V. S.D. contributed to genetically inferred ancestry prediction from WES data with supervision from S.J.S.. M.C. designed and performed all the rare variant analyses with supervision from D.H.G., S.J.S., and J.M.F., and statistical support from R.M.C. and D.E.A.. M.C. and D.H.G. wrote the first draft of the manuscript with contributions from D.S.G., A.O.F.C., M.E.K., and J.K.L., and feedback from all authors. M.C. and D.H.G. curated the following and final versions of the manuscript.

## Methods

### Genetics of Neurodevelopment in African Americans (GENAA) study participants

#### Recruitment

Our Autism Center of Excellence (ACE) Network^2,3^ includes four areas across the country contributing to the GENAA Consortium that recruit and assess families who self-identify as Black or African American (AfrAm) and include individuals with known or suspected autism: Albert Einstein College of Medicine (Bronx, NY), Emory University (Atlanta, GA), Washington University (WUSTL; St. Louis, MO), and UCLA (Los Angeles, CA). Families are recruited in GENAA through public resource fairs for families with developmental disabilities, social media, word-of-mouth, ads in public areas such as mass transit stops, clinic waitlists, and clinician referrals. Family eligibility criteria include: (1) at least one AfrAm offspring individual with known or suspected autism spectrum disorder (ASD); (2) absence of other known neurological or genetic disorders in the proband; (3) participation from at least one biological parent. Potential nonautistic siblings are also offered to participate in the study. As of August 2025, our Network has recruited 1,677 families with at least one AfrAm offspring individual with confirmed ASD diagnosis; a total of 4,634 individuals, including 1,912 autistic offspring individuals, participate in our research program and have contributed a biomaterial for sequencing.

Probands are initially screened (mostly by phone interview with a parent) for the presence of ASD-related symptoms using the Social Communication Questionnaire (SCQ)^59^ or the Modified Checklist for Autism in Toddlers, Revised (M-CHAT-R)^93^ (Supplementary Table 11).

Informed consent was obtained from the parent or legal guardian of all participants in this study, as well as either consent or assent from participating minors and young adults where age- and developmentally appropriate. This study was approved by the Institutional Review Boards at each recruitment site.

Recruitment and data collection for the participants described in this paper (Supplementary Table 1,2) took place from 2013 to 2022, with additional families recruited at WUSTL from 2008 to 2013.

#### Behavioral assessment

Timing of behavioral assessment varied based on participant phenotype and age. Individuals enrolled at age 36 months through adulthood with known or suspected ASD were assessed at a single time point (“school-age cohort”) at all four recruitment sites. At the Emory and WUSTL sites, individuals enrolled at age 18-35 months with known or suspected ASD were assessed at enrollment (“Time 1”) and again ≥ 18 months later (“Time 2”). Participating parents and siblings not suspected of having ASD were assessed at a single time point.

Briefly, each participant was assessed for cognition (Differential Ability Scales-II or DAS-II^94^, Raven’s Progressive Matrices^95^, Peabody Picture Vocabulary Test^96^, Mullen Scales of Early Learning or MSEL^97^), quantitative autistic traits such as social responsiveness (Social Responsiveness Scale or SRS^63^) and video-referenced Reciprocal Social Behavior or vrRSB^98^ (parent-, spouse- or self-report), and handedness (adapted from Edinburgh Handedness Inventory^99^). A clinical best estimate ASD diagnosis for the proband was confirmed or made according to whether criteria for ASD were met on a DSM-5 (Diagnostic and Statistical Manual of Mental Disorders, 5^th^ edition) checklist based on the integration of results and information from the forementioned assessments, the Autism Diagnostic Observation Schedule 2 (ADOS-2)^100,101^, the caregiver DSM-5 interview^102^, and a caregiver Autism Diagnostic Interview-Revised (ADI-R)^103^ if necessary. Information on the proband’s medical history, family medical history, and proband’s skills of daily living (Vineland Adaptive Behavior Scales or VABS^104^, Child Behavior Check List or CBCL^105^) were obtained from parent interviews. The primary caregiver of each proband was also interviewed to document challenges and obstacles experienced in obtaining an accurate ASD diagnosis and services (through the Event History Calendar Interview or EHCI^2^) and parents were asked to describe any services received, including duration and perceived efficacy.

Select sites offered remote assessments conducted through phone and video appointments, beginning during the COVID-19 pandemic. Remote assessments used a modification of the Childhood Autism Rating Scale (CARS-2obs)^106^ in place of the ADOS-2, with additional information gathered from the parent as needed for diagnostic certainty using the ADI-R. Cognition was assessed using the Raven’s-2 or Raven’s Standard/Colored (as age- and developmentally appropriate) in place of the DAS-II.

Supplementary Table 11 lists the assessment battery for each participant group.

Age of participants was retrieved for analysis at time of most recent behavioral assessment when available, otherwise at time of biomaterial collection.

#### Biomaterial collection

Participants of the GENAA cohort donated a biomaterial in the form of whole blood (venipuncture) or saliva (Oragene OGR-500 or OGR-575; DNA GenoTek, Ontario, Canada). EDTA-stabilized whole blood DNA was extracted by the National Institute of Mental Health (NIMH) Repository & Genomics Resource (or NRGR, currently known as Sampled, Piscataway, NJ) using standard protocols. DNA from saliva was extracted according to the manufacturer protocol (prepIT.L2P; DNA GenoTek, Ontario, Canada) and quantified by fluorimetric assay (Quant-iT dsDNA Broad Range Assay Kit, Invitrogen, USA).

#### Additional biomaterials from the Autism Genetic Resource Exchange (AGRE) cohort

This GENAA study also includes additional individuals of AFR descent recruited as part of the AGRE cohort^107^ (genotyping dataset after quality control [QC] – total: n = 1,060; of AFR GIA: n = 145; whole-exome sequencing [WES] dataset after single nucleotide variant [SNV] QC – total: n = 179; of AFR GIA: n = 150; Supplementary Table 1,2). Their purified DNA was obtained from the NIMH Repository & Genomics Resource and was derived from whole blood, where available, as described above. DNA from lymphoblastoid cell lines (LCLs) was used for individuals for whom whole blood DNA was not available. AGRE and GENAA individuals were combined for all analyses except from genetically inferred ancestry (GIA) prediction on genotyping data. Throughout this manuscript, GENAA mostly refers to GENAA and AGRE combined. Most genotyped AGRE individuals were labeled as of European (EUR) GIA.

### Genetic data generation

#### Genotyping

Genotyping was performed by the UCLA NeuroGenomics Core (UNGC) in multiple batches using the Illumina platform with standard manufacturer protocols. Array types included Omni-1 (n = 1,151), Omni-2.5 (n = 978), and Global Screening Array (n = 50). Supplementary Table 1 breaks down the number of families and individuals with genotyping data, before and after QC.

#### WES

WES was performed by the UNGC or the Genomics Core of the Institute of Human Genetics (IHG) at UCSF in four batches using standard manufacturer protocols. Sequencing libraries were prepared using the KAPA HyperPrep kit for library preparation (Roche) in conjunction with the Nimblegen SeqCap EZ v2.0 or HyperExome exome enrichment kits (Roche) according to manufacturer protocols. Supplementary Table 1 breaks down the number of families and individuals with WES data, before and after QC.

Before QC, there were 776 GENAA individuals with matched genotyping and WES data. After QC, these became 760.

### Genotyping data processing and QC

#### Initial per-batch QC

Quality filtering was performed on each genotyping batch separately using PLINK v1.07 or later. Within-batch family relationships were validated using PLINK identity-by-descent (IBD) estimation (--genome). Expected sex was validated for all individuals using PLINK --check-sex and results were used as supporting evidence for mislabeled samples. Data were filtered to exclude individuals or markers with ≥ 2% missingness and markers with Hardy-Weinberg equilibrium (HWE) *P* < 10^-7^.

#### Imputation

Genomic imputation was run with the TOPMed Imputation server^108^. Before uploading to the TOPMed server, the following pre-imputation QC steps were run. Duplicate single nucleotide polymorphisms (SNPs) were removed, as well as those not containing an A, T, G, or C, using PLINK2 v2.00a2^54^. Individuals with a call rate < 0.9 were also removed. The data was then compared to the TOPMed reference panel using McCarthy Group Tools (HRC-1000G-check-bim-v4.3.0). The toolkit checks the strand, alleles, and positions against the reference data. Then, it removes palindromic SNPs with a minor allele frequency (MAF) > 0.4, SNPs with an allele frequency with > 20% difference from the reference panel, and alleles that are different from the reference. The imputation was run using TOPMed-r2 reference panel, Eagle v2.4 phasing, an r^2^ filter of 0.3, and in GRCh38/hg38 genome build^108,109^. The imputation server removes indels, monomorphic sites, SNPs with an allele mismatch from the reference panel, and SNPs with a call rate < 0.9. After imputation, SNPs with r^2^ > 0.8 were kept, which left a final set of 55,254,922 common variants. Prior to analysis, these SNPs were selected by MAF > 0.01 and HWE *P* > 10^-6^.

#### Post-imputation sample QC

Cross-batch genetic relationships in GENAA were verified with IBD in KING v2.3.2^110^. One individual from each twin pair was removed from analysis. Finally, genetic sex was re-verified using PLINK v1.90b6.24. In addition, only individuals with high-quality phenotype annotation were kept for analysis. This left a final total set of 2,131 individuals who pass QC in the GENAA genotyping dataset (Supplementary Table 1,2).

#### Integration with the Simons Foundation Powering Autism Research for Knowledge (SPARK) cohort genotype data

SPARK is the largest ASD research study to date^12^. The SPARK dataset SPARK Consortium iWES v1.1^111^ (based on WES and Global Screening Array version 2) was integrated with GENAA for the common variant analyses. SPARK individuals who also participated in the Simons Searchlight cohort, previously named Simons Variation in Individuals Project (Simons VIP), were removed since the cohort specifically recruited carriers of 16p11.2 and 1q21.1 deletions or duplications. Genotype imputation and pre- and post-imputation QC were performed as described for the GENAA data. In addition, SPARK imputed genotype data were filtered for genotype call rate > 0.95, MAF > 0.01, HWE *P* > 10^-6^, and non-duplicate SNPs using PLINK 2.0. SPARK individual phenotypic information for downstream analysis (ASD status and SQC raw total score) was extracted from the SPARK phenotypic data release SPARK Collection Version 7, from December 2021^111^.

### Genetic ancestry inference

#### GIA

Throughout this manuscript, we discuss genetic descent of GENAA and other cohorts’ participants using the previously described term “GIA”^51^, which refers to the genetic characterization of individuals within a group who likely share recent biological ancestors as inferred by a method of choice (inference method and reference data). We use this term to emphasize its distinction from instances of genetic ancestry, such as the African continent populations in reference panels (with ancestors from Africa), for which information about the origin of the recent biological ancestors is known^51^. Additionally, this term is appropriate for the description of admixed populations like AfrAms in the United States^41,42,52,53^. Therefore, in the context of this work, “AFR GIA” refers to an admixed group of individuals in the United States who have recent biological ancestors inferred to be of African ancestry and thus show partial or total ancestry originating from Africa. Similarly, “EUR GIA” refers to individuals with recent biological ancestors inferred to be of European ancestry and, therefore, partial or total ancestry originating from continental Europe.

GIA was calculated in both imputed datasets, GENAA and SPARK, separately using a clustering method based on principal component analysis (PCA)^112^. GIA labels were assigned to the GENAA and SPARK individuals by genetic similarity to the labeled reference populations of the 1000 Genomes Project^36,113,114^. Briefly, SNPs were first subset to those present in the 1000 Genomes Project dataset: 426,243 SNPs were left for GENAA and 4,000,344 SNPs were left for SPARK. The data was then merged with the 1000 Genomes Project data representing individuals from the five “superpopulations”: AFR, AMR (Admixed American), EAS (East Asian), EUR, and SAS (South Asian). The combined data of individuals was then mapped onto the PCs from the 1000 Genomes Project reference populations using bed_projectPCA, a function in the R^115^ package bigspnr v1.12.4^116^. The centers of each reference population were identified, in PCA space, and the fixation index (FST) between those populations as well as the Euclidean distance between centers were calculated for each pair of superpopulations^30^. For every sample, the Euclidean distance to each population center was calculated. If the closest population center to an individual was either AMR or AFR, they were assigned to that corresponding GIA group. For the remaining populations, a square distance threshold was used to determine the corresponding GIA label. The calculation is outlined in the reference^30^. Individuals whose minimum distance to a population center was less than this threshold were assigned to that GIA group, while those not passing this threshold for any population were not assigned to any GIA group (**Supplementary Fig. 1**). This resulted in final AFR GIA groups of 1,060 total individuals for GENAA and 3,766 for SPARK (Supplementary Table 3).

#### Local ancestry inference

Local ancestry was inferred using RFMix v2.03-r0^65^. An EUR reference panel (CEU or Utah Residents with Northern and Western European ancestry continental population) and an AFR reference panel (YRI or Yoruba in Ibadan Nigeria continental population) from the 1000 Genomes Project catalog were used to identify the ancestry partial subsets^113,114^.

The samples were first joined with all the 1000 Genomes Project reference populations and, after joint PCA, run with admix-kit “select-admix-individuals” to select admixed genomes with significant AFR and EUR GIA^66^. The following parameters were used: --superpop1 EUR --superpop2 AFR - -exclude-pop2 ASW,ACB; the ASW or African Ancestry in Southwest US and ACB or African Caribbean in Barbados reference individuals were excluded from the AFR “superpopulation” given their admixture. Briefly, the software runs a joint PCA analysis of the provided data with the reference populations from the 1000 Genomes Project to identify which defined continental population individuals are most similar to. Admixed individuals are identified as those that lie between the ancestral populations of interest based on the PCs, in this case AFR and EUR. Then, the joint population was subset to the individuals of interest assigned to AFR, EUR, or admixed AFR-EUR inferred ancestries. Finally, these samples were run by chromosome in RFMix to determine local ancestry based on genetic similarity to the CEU and YRI reference data. RFMix divides individual’s chromosomes into genomic windows and compares those windows to reference data to determine which defined continental population is most like each window. It produces ancestry estimates for each window as well as a global diploid ancestry estimate for each of the 22 autosomal chromosomes. This global chromosome ancestry estimate was averaged over all autosomes to get AFR and EUR global genetic ancestry proportions for all individuals in the analysis.

### Analysis of common variation

#### ASD diagnosis genome-wide association study (GWAS)

ASD diagnosis categorical GWASs for the GENAA and SPARK cohorts were performed with PLINK2 v2.00a2^54^. An unrelated population of individuals of AFR GIA was created from the GIA estimates. Cases were chosen from the probands, and controls were taken from the parents of individuals not included in the case group. For GENAA, the final unrelated AFR GIA group had 272 cases and 268 controls (Supplementary Table 4). The final SPARK group had 1,127 cases and 1,014 controls (Supplementary Table 4). The results from GENAA and SPARK were meta-analyzed using METAL with inverse variance-based weighting^117^. Variants with MAF > 0.01 and HWE *P* > 10^-12^ were selected for analysis. The GWAS was run with PLINK2 as a logistic model. The covariate variances were standardized and the first ten genotype PCs were used as covariates.

In this and subsequent GWASs, the Bonferroni threshold for genome-wide significance of *P* ≤ 5.0 x 10^-8^ was used to define significant SNP associations and the *P* ≤ 10^-5^ threshold was considered suggestive of association. In addition, results were visualized with a julia package, *GeneticsMakie.jl*^118^, and quantile-quantile (Q-Q) plots were used for QC.

Spearman correlation was computed between β coefficients of selected SNPs from the published EUR-based ASD diagnosis GWAS^9^ and the AFR GIA based ASD GWAS GENAA and SPARK meta-analysis. Lead SNPs to compare between the two studies for their effect sizes were selected from the EUR GWAS as having *P* < 10^-5^ and being independent from each other at R^2^ < 0.1 based on linkage disequilibrium (LD) pruning.

Of the 12 ASD-associated SNPs from the published EUR-based ASD diagnosis GWAS^9^, only rs325506 was among these defined lead SNPs and included in the correlation analysis. The other 11 SNPs were excluded from analysis for different reasons, as follows. Two SNPs (rs201910565 and rs71190156) were not part of the AFR GIA based GWAS meta-analysis of GENAA and SPARK as they could not be imputed from their genomic coordinates. SNP rs111931861 was excluded since having imputation information score < 0.9 in the EUR-based ASD GWAS summary statistics file. Three SNPs (rs1620977, rs10149470, and rs16854048) were removed as they had *P* > 10^-5^ in the EUR-based ASD GWAS summary statistics file. Finally, five SNPs (rs910805, rs10099100, rs2388334, rs11787216, and rs1452075) had *P* < 10^-5^ in the EUR-based ASD GWAS summary statistics file but were not independent from each other at R^2^ < 0.1 based on LD pruning: these non-lead SNPs were excluded from the correlation analysis but displayed and labeled in Fig. 1D for reference.

In addition, Spearman correlation was computed between β coefficients of selected SNPs from the two AFR GIA based ASD GWASs in GENAA and SPARK. SNPs from both analyses were ranked by their p-values and the top one million SNPs in common were selected for LD pruning. Independent SNPs were defined at R^2^ < 0.1 and ranked by their p-values first in GENAA and then in SPARK. The top 100 independent SNPs in common were selected for correlation analysis.

β coefficients for the known EUR-derived ASD-associated common SNP rs1452075 C>T from the EUR-based ASD GWAS and the AFR GIA based ASD GWAS meta-analysis in GENAA and SPARK were tested for heterogeneity through the Cochran’s Q test. This test assesses the null hypothesis of homogeneous effect sizes across studies and therefore, a p-value > 0.05 was considered indicative of consistent effect sizes. Two heterogeneity statistics were reported: τ^2^ (between-study variance or variance of the underlying distribution of true effect sizes) and *I*^2^ (proportion of variability in effect sizes due to true heterogeneity and not chance).

Local ancestry was analyzed for the known EUR-derived ASD-associated common SNP rs1452075 C>T in AFR GIA individuals, both total and just autistic, of the GENAA and SPARK cohorts used in the ASD GWASs. Briefly, we used the local ancestry predictions, derived from genotyping data through RFMix as already described, to count both the autistic individuals and the total individuals included in the GENAA and SPARK AFR GIA based ASD GWASs who had: (1) zero alternate allele (T) copies, one alternate allele copy, or two alternate allele copies for the SNP on AFR background; and (2) AFR/EUR, AFR/AFR, or EUR/EUR diploid local ancestry at the genomic segment encompassing rs1452075 (Supplementary Table 18). The observed individual local ancestry counts were then compared to expected ones, based on individuals’ proportions of AFR global genetic ancestry, under the HWE. For each individual *i*, the expected probabilities of observing homozygous AFR local ancestry, heterozygous AFR/EUR local ancestry, and homozygous EUR local ancestry at this locus were computed as p*_i_*^2^, 2 x p*_i_* x (1 - p*_i_*), and (1 - p*_i_*)^2^, respectively, with p*_i_* representing the individual proportion of AFR global genetic ancestry and (1 - p*_i_*) representing the individual proportion of EUR global genetic ancestry (based on the two-way admixture model used for ancestry deconvolution with RFMix). These values were then summed across individuals to get the expected individual counts for comparison. Two-sided Chi-squared tests were used to identify potential deviations of the observed counts from the expected counts, and p-values < 0.05 were considered significant.

#### SAIGE ASD diagnosis GWAS

To increase sample size and power, SAIGE^55^ was used to run a GWAS for ASD diagnosis in the individuals, parents and offspring, with AFR GIA in both GENAA and SPARK. SAIGE calculates and incorporates a genetic relatedness matrix to account for relatedness in a sample population^55^. This allows for related individuals to be included in the analysis and increases the sample size available for the GWAS (459 cases and 601 controls from GENAA and 1,931 cases and 1,833 controls from SPARK) (Supplementary Table 4). ASD diagnosis was run as the trait of interest with age, sex, and the first ten genetic PCs used as covariates. The results from these analyses were meta-analyzed with METAL as previously described^117^.

Spearman correlation was computed between β coefficients of selected SNPs from the published EUR-based ASD diagnosis GWAS^9^ and the SAIGE AFR GIA based ASD GWAS GENAA and SPARK meta-analysis. Lead SNPs to compare between the two studies for their effect sizes were selected from the EUR-based ASD GWAS as having *P* < 10^-5^ and being independent from each other at R^2^ < 0.1 based on LD pruning.

#### GWASs for quantitative autistic traits: SCQ and SRS

Additional AFR GIA based GWASs for the quantitative autistic traits captured by the assessment tools SCQ^59^ and SRS^63^ were performed using GENAA combined with SPARK and GENAA combined with the Adolescent Brain Cognitive Development (ABCD) cohort^61,62^, respectively.

Briefly, these were the steps performed on the ABCD data. Genotyping (Affymetrix NIDA Smokescreen Array) and behavioral (SRS) data were obtained from the ABCD 4.0 National Data Archive data release.

GIA for the ABCD individuals was inferred as follows. First individuals whose genotype data was identified as low quality by ABCD were removed. SNP genotype data were then filtered for individual call rate (> 0.90), genotype call rate (> 0.95), MAF (> 0.01), HWE p-value (> 10^-7^), non-duplicate SNPs, and non-palindromic SNPs using PLINK 2.0 and 1.9, leaving a final set of 333,158 common SNPs passing QC. GIA by genetic similarity was determined by merging to the 1,000 Genomes Project genotypes, followed by PCA, and application of a k-nearest neighbors classification algorithm to the first ten genotype PCs.

Then, for each defined GIA group, a genetic relationship matrix was computed from autosomal chromosomes using GCTA^60^ v1.93. Imputation of the ABCD genotypes was performed exactly as already described for GENAA and SPARK. Only high imputation quality SNPs (r^2^ > 0.8) were kept for downstream analysis. As done in SPARK, after imputation, for the AFR GIA group defined in ABCD, SNPs were further filtered for genotype call rate (> 0.95), MAF (> 0.01), HWE p-value (> 10^-7^), and non-duplicate SNPs using PLINK 2.0.

SCQ data were available for GENAA and SPARK only, whereas SRS data were available for GENAA and ABCD only. In SPARK, the SCQ Lifetime version was completed by a parent/adult or other primary caregiver for autistic participants and their nonautistic siblings aged 2-18 years. For SPARK autistic participants, SCQ raw total scores were selected for GWAS and standardized with a rank-based inverse normal transformation. The ABCD study distributed a shortened version of the SRS, consisting of 11 Likert-style items. The SRS raw total scores were selected for GWAS and standardized with a rank-based inverse normal transformation. After QC, 321 ASD cases (259 males, 62 females) in the GENAA cohort, and 1,278 ASD cases (1,029 males, 249 females) in the SPARK cohort were included in the SCQ GWAS analysis, and 407 ASD cases (334 males, 73 females) in the GENAA cohort, and 1,953 individuals (976 males, 977 females) in the ABCD cohort were included in the SRS GWAS analysis (Supplementary Table 4). Genome-wide mixed linear model association analysis for each quantitative autistic trait measure within each cohort was run using mlma in GCTA^60^, adjusting for age in months, sex, and the first 10 genotype PCs. For each quantitative autistic trait measure, GWAS meta-analysis combining cohorts was performed in METAL using inverse variance-based weighting^117^.

#### ASD polygenic scoring and analysis

ASD polygenic scores (PGSs) were calculated for individuals in the GENAA and SPARK cohorts (Supplementary Table 4) as an estimate of an individual’s predisposition to ASD, using EUR-derived polygenic summary statistics for 35,087 variants^9^. The score was accessed through the PGS Catalog (PGS000327), an online repository of published PGSs^67,68^. Scores were downloaded in GRCh37 genome build format and converted from GRCh37 to GRCh38 to match PGS variant coordinates to GENAA and SPARK imputed variant call file SNP coordinates. Scores were converted and calculated using pgsc_calc v0.5.3, the PGS calculator published by the PGS Catalog. PGSs were then root mean square scaled by GIA and cohort (GENAA, SPARK, and Combined) and centered on zero. ASD PGSs were compared between autistic and nonautistic offspring by GIA and cohort (GENAA, SPARK, and Combined) using Welch’s t-tests to account for unequal variances and sample sizes. Moreover, to combat the effects of class imbalance in our data, frequency weights derived from case/control ratios were calculated and incorporated into a weighted logistic generalized linear model (GLM) when estimating the association between ASD PGS (root-mean-square-scaled) and diagnosis (Supplementary Table 19). Cohort (GENAA or SPARK) was used as covariate in the weighted logistic GLMs. Coefficients from the weighted logistic GLMs were exponentiated and reported as odds ratios (ORs). Significance was set at the 0.05 level.

#### Polygenic transmission disequilibrium testing (pTDT)

Polygenic transmission disequilibrium testing^8^ was performed using the Python tool pTDT v1.0.0 (Supplementary Table 4,19). In brief, PGSs available for both parents in a family were averaged to obtain a mid-parent score for each offspring individual. The deviation of the observed offspring individual PGS from the mid-parent score was then measured to examine potential over transmission. For interpretation purposes, the raw deviation score was standardized by dividing it by the standard deviation of the mid-parent PGS distribution in the study population. A pTDT deviation of 0 implies that the offspring individual’s PGS matches the mid-parent score, indicating no unusual transmission of contributing alleles. A deviation significantly above 0 indicates an over-transmission of contributing alleles to the offspring individual. To assess if mean pTDT deviation was significantly different than 0, the test statistic was evaluated with two-sided one-sample t-tests. P-values < 0.05 were considered significant. All PGSs were root mean squared scaled by cohort (GENAA, SPARK, and Combined) and GIA prior to assessing polygenic transmission disequilibrium.

#### Ancestry partial polygenic scoring for ASD

The output from RFMix^65^ was run with the admix-kit toolkit to calculate ancestry partial polygenic scores^66^. The datasets were subset to individuals, both parents and offspring, with AFR, EUR, or admixed AFR-EUR genomes from GENAA and SPARK (Supplementary Table 4). The same EUR-derived ASD PGS was applied to both the AFR and EUR genome portions separately^9,66^. Individuals had resulting AFR partial, EUR partial, and total ASD PGSs (Supplementary Table 20). A logistic GLM was fit for ASD diagnosis for each of the AFR partial, EUR partial, and total scores, using the R function glm() in the stats v3.6.2 R^115^ package. Using the global genetic ancestry proportion estimates, individuals were also binned by their proportion of AFR ancestry, and logistic models were fit to the ancestry partial and total scores within these bins. As in PGS analysis, outlined above, frequency weights were calculated by AFR global genetic ancestry proportion bins and applied to the models. ASD ORs and their 95% confidence intervals (CIs) were calculated for each of the GLMs and used for comparison. For the ancestry partial scores, age, sex, and cohort (GENAA or SPARK) for the combined population, were used as covariates. The enrichment for two-parent families of offspring with medium proportion of AFR global genetic ancestry including one AFR GIA and one EUR GIA parents within all two-parent families of offspring part of the ancestry partial ASD PGS analysis was tested with two-sided Fisher’s exact test in GENAA (Supplementary Table 20).

#### WES data processing, QC, and analysis

WES data was generated and analyzed for a total of 1,324 individuals in the GENAA cohort (Supplementary Table 1,2). After initial QC of FASTQ files using FastQC (version 0.11.9) and MultiQC (version 1.11), data processing was run on Terra (https://app.terra.bio/), a Google Cloud hosted platform for biomedical data analysis and genomic research collaboration operated by the Broad Institute Data Sciences Platform team. Processing was performed using specific cloud-optimized Workflow Description Language (WDL)^119^ scripts and WDL Analysis Research Pipelines (WARP)^120^ (Data Sciences Platform, Broad Institute. (2022, February 9) *warp-pipelines/Exome-Analysis-Pipeline* [workspace] Retrieved June 23, 2022, https://app.terra.bio/#workspaces/warp-pipelines/Exome-Analysis-Pipeline) curated and shared by the Broad Institute Data Sciences Platform.

First, paired FASTQ files were converted to read group specific unmapped BAM files. Then, single-sample processing steps for germline SNV and small Indel discovery in human WES data were run through the ExomeGermlineSingleSample_v3.0.3 WARP. This pipeline performs QC at various steps, alignment to the hg38 reference genome, duplicate marking, base recalibration, and variant calling according to the Genome Analysis Toolkit (GATK) best practices and adhering to the Functional Equivalence specification^121^. It generates multiple per-sample final outputs, including QC metrics, an aligned CRAM file, and a GVCF file. The pipeline was run using exome enrichment kit specific genomic intervals. Additionally, a patch version of the VariantCalling_v2.0.3 workflow was used to generate non-reblocked GVCFs, offering higher genotype quality resolution for *de novo* variant calling, in place of the workflow default reblocked GVCFs. At this stage, samples that were extreme outliers for QC metrics collected through Picard or that were flagged as potentially contaminated (VerifyBamID FREEMIX > 0.02) were excluded from downstream steps. Starting from the non-reblocked GVCFs, joint genotyping, variant quality score recalibration, and post-calling variant QC were run on two sample “super batches” defined by exome enrichment kit with the JointGenotyping_v1.5.2 workflow using kit specific genomic intervals. These runs generated multiple per-batch variant calling QC metrics and two multi-sample filtered site annotated VCF files. The two multi-sample VCFs were used as inputs for standard QC confirming sample identity (expected biological sex and familial relationships) through PLINK v1.90b and for downstream SNV/Indel analysis steps.

Starting from the per-sample WES-derived CRAMs, additional germline copy number variant (CNV) calling was performed on Terra using the GATK-gCNV workflow as previously described^13,122^. Prior to CNV processing, samples were clustered via PCA into batches by read-depth profile similarity over a set of predefined and filtered genomic intervals differentiating common exome enrichment kits^13,122^. The GENAA WES samples were clustered into three “super batches” via a primary PCA. A secondary PCA further subdivided one of the “super batches” into four clusters, two of which contained too few samples (< 40) to accurately run the GATK-gCNV unified probabilistic model^13,122^. For this reason, 44 GENAA WES samples were not CNV processed. GATK-gCNV cohort mode was run for each of the four final clusters, using a maximum of 200 samples per cluster. For two clusters with > 200 samples, GATK-gCNV case mode was run on the remaining samples with corresponding cohort mode pretrained models. The CNV events were queried over curated non-overlapping intervals corresponding to all exons of canonical transcripts of protein-coding genes from the GENCODE v33 annotation that were further defined as well-captured in all samples, as previously described^13,122^. After per-sample processing, the non-reference copy number calls of each sample with quality score (QS) ≥ 20 were considered for defragmentation^13,122^. Then, concatenated calls from samples of all clusters were compiled into a single table and merged into unique sites by ≥ 80% reciprocal overlap^13,122^. Site frequencies (SFs) were assigned to each site as the proportion of GENAA WES CNV processed samples that had the site in the combined dataset. For each CNV, number of spanned well-captured intervals and exons were calculated for further downstream filtering^13,122^.

#### De novo SNV/Indel calling and annotation

Variant calling was performed on IBD-verified trio families consistent with prior SPARK and Autism Sequencing Consortium (ASC) analyses to ensure comparability^11,13^. Variants were processed using Hail (https://github.com/hail-is/hail) de_novo() function with the non-neuro subset of the Genome Aggregation Database (gnomAD) GRCh38 exomes v2.1.1 (gs://gnomad-public/release/2.1.1/liftover_grch38/ht/exomes/gnomad.exomes.r2.1.1.sites.liftove r_grch38.ht) as priors. Putative *de novo* variants were dropped if they had a genotype quality (GQ) score < 25, were present within this gnomAD subset at a frequency > 0.01%, or present in the GENAA cohort at a frequency > 0.1%. Further variants were dropped if “ExcessHet” was in the ‘Filters’ field, if the progeny’s allele balance was < 0.3, or if the depth ratio (offspring individual read depth divided by the sum of parental read depth) was < 0.3. For indels only, variants were dropped if the probability of their allele balance (based on a binomial distribution centred around 0.5) was < 0.05. Variants were annotated using the Variant Effect Predictor (VEP)^123^. Read-level data from the offspring and both parents were inspected for all putative *de novo* indels using the Integrative Genomics Viewer (IGV)^124^ to exclude inherited indels represented differently in the VCF in a parent. Only autosomal *de novo* SNVs/Indels were selected for downstream analysis.

#### Rare CNV filtering and annotation

After variant and sample QC on the CNV call set, high-quality rare CNV calls were defined as those with SF < 1%, spanning > 2 well-captured exons, and passing specific CNV type and interval dependent QS thresholds, as previously described^13^. Three GENAA WES CNV processed samples were identified as outliers^13^ and excluded from downstream CNV analysis.

Several annotations were added to the CNV call set. A deletion was annotated as overlapping a gene if it covered ≥ 10% of the non-redundant exon base pairs of the gene, whereas a duplication was annotated as overlapping a gene if it covered ≥ 75% of the non-redundant exon base pairs of the gene^13^. In addition, CNVs were annotated against gnomAD v4.1.0 (matched CNV type and ≥ 80% reciprocal overlap) with their genetic ancestry group specific SFs and the maximum SF value observed across genetic ancestry groups in gnomAD (or gnomAD grpmaxSF) was selected as the most appropriate annotation for further frequency-based call set refinement. CNVs with gnomAD grpmaxSF ≤ 1% or not found in gnomAD (gnomAD grpmaxSF = NA) were defined as rare (truly rare in any genetic ancestry group, even in the group in which they were observed most frequently). Moreover, with the goal of highlighting potentially novel ASD association signals, rare CNVs were annotated against a curated list of CNV loci (matched CNV type and ≥ 50% reciprocal overlap) as being ASD-associated and/or genomic disorder and/or autism-associated (GDA-associated) events (Supplementary Table 25). The list of CNV loci underlying this rare CNV classification was curated as follows. We gathered a catalog of 152 GDA-associated CNVs of interest, composed of 121 genomic disorder (GD) loci from gnomAD v4.1.0 and of 31 ASD-associated loci from the Simons Foundation Autism Research Initiative (SFARI) Gene database (2024 Q3 release). The GD sub list includes recurrent CNVs associated with syndromic features, but not necessarily with ASD or other neurodevelopmental disorders (NDDs), that have well-defined breakpoints (and therefore, coordinates) since mainly due to nonallelic homologous recombination events^13^. On the other hand, the ASD sub list from SFARI consists of CNV loci that have been significantly linked to ASD but show less consistent coordinates that represent the broader genomic range in which the events have been observed in carriers from various ASD studies (https://gene.sfari.org/user-guide/cnv/). For these reasons, when curating the final 152-CNV locus union list, the two coordinate systems were kept independent (no site merging) but an annotation was added indicating which recurrent GD CNVs overlapped an ASD-associated locus (or SFARI locus) and could, therefore, be considered known genetic factors for ASD. In particular, a GD CNV was defined as confidently overlapping a SFARI CNV (GD-SFARI CNV) if (1) the two sites showed the same CNV type and (2a) ≥ 50% reciprocal overlap or (2b) ≥ 80% nonreciprocal overlap (one site almost entirely spanned or was spanned by the other). This resulted in the definition of 19 GD CNVs out of 121 as specifically ASD-associated (and, in parallel, of 17 SFARI CNVs out of 31 as recurrent GD loci). Based on this curated list, if a rare CNV confidently overlapped any of the 152 loci, it was annotated as GDA-associated, whereas if it overlapped any of the 31 SFARI loci, or 19 GD-SFARI loci, or both, it was annotated as ASD-associated. Only autosomal rare CNVs were selected for downstream analysis.

#### Integration of GENAA rare variants with variant sets from the ASC, SPARK, and the Simons Simplex Collection (SSC) cohorts

To improve power, comparability, and generalizability of rare variant analyses, the GENAA WES rare variants were combined with variant sets from the largest study aggregating exome-sequenced samples from three autism cohorts, ASC, SPARK, and SSC^13^ (Supplementary Table 21). Many processing steps for the GENAA WES variants and samples were aligned with those from this seminal study^13^. Specifically, 35,257 *de novo* SNVs/Indels from 20,526 fully phase-able offspring individuals (autistic: n = 15,034; nonautistic: n = 5,492) from ASC, SPARK, and SSC^13^ were integrated with GENAA. In addition, rare CNVs from ASC, SPARK, and SSC^13^ were annotated with gnomAD v4.1.0 grpmaxSF and further filtered and classified as described for GENAA. For combined CNV analysis, 20,638 rare CNVs from 18,048 fully phase-able offspring individuals (autistic: n = 13,182; nonautistic: n = 4,866) from ASC, SPARK, and SSC^13^ were integrated with GENAA. Two autistic fully phase-able offspring individuals were part of both GENAA and ASC, and therefore, included only once in analysis, as GENAA individuals.

The GENAA WES dataset included nine pairs of monozygotic twins. Only one twin for each pair was kept for analysis. When possible, autism diagnosis, whole blood DNA, and carrier status were preferred during selection; otherwise, twin selection was random. After technical and identity QC, removal of one twin per pair, and selection of only individuals with high-quality phenotype annotation, there were 1,297 GENAA individuals who passed SNV QC and 1,253 individuals passing CNV QC (Supplementary Table 1,2). A total of 434 *de novo* SNVs/Indels (409 SNVs and 25 Indels) from 316 fully phase-able offspring individuals (autistic: n = 248; nonautistic: n = 68) from GENAA were analyzed in combination with the other ASD cohorts, whereas 950 rare CNVs from 701 GENAA offspring individuals (autistic n = 545; nonautistic n = 156) were considered and combined for analysis (Supplementary Table 21). Family offspring from all ASD cohorts (GENAA, ASC, SPARK, and SSC) were further grouped by their GIA (AFR or EUR), defined by genetic similarity to labeled reference populations using a uniformed protocol (see section below) (Supplementary Table 21).

Rare variants were further annotated and categorized for analysis, consistently across cohorts, as follows. *De novo* missense SNVs were annotated with their corresponding Missense badness, PolyPhen-2, and Constraint (MPC) score^70^ using Hail and categorized, as previously described^13^, as MisB (MPC ≥ 2), MisA (MPC ≥ 1 and MPC < 2), and of predicted low impact (Mis MPC < 1). Genes impacted by *de novo* protein-truncating variants or PTVs (defined as frameshift, splice site acceptor and donor, and stop gain variants) and rare CNVs were annotated for constraint through their corresponding loss-of-function observed/expected upper bound fraction (LOEUF) score and LOEUF score decile^69^. The gene constraint annotations were retrieved from the publicly available file “gnomad.v2.1.1.lof_metrics.by_gene.txt”^69^, merging by gene name, or by “gene_id” if available. The longest gene version was kept for duplicated gene name entries in the table. Rare CNVs spanning multiple genes were annotated with the LOEUF score of the most constrained impacted gene (therefore, with the lowest score among those available). A hard LOEUF score threshold of 0.4 was used to define constrained genes (LOEUF < 0.4). Impacted genes were further categorized for their constraint into four groups, as having LOEUF score in the 1^st^, 2^nd^, 3^rd^, or 4^th^-10^th^ deciles^13^, to create PTV, deletion, and duplication sets for downstream testing. In addition, rare CNVs were divided into four testing groups by their size: > 1Mb, [1Mb, 100kb), [100kb, 10kb), and ≤ 10kb.

#### Genetic ancestry inference

GIA was inferred for the exome-sequenced individuals from the four cohorts with a uniformed protocol as follows. Labeled reference populations from the 1000 Genomes Project (including AFR, AMR, EAS, FIN or Finnish, NFE or Non-Finnish European, OTH or Other, and SAS) and the Human Genome Diversity Project or HGDP (including AFR, AMR, EAS, MID or Middle Eastern, NFE, OTH, SAS) (n = 4,119^36,113^) were used as training dataset for a random forest classifier, with the exception of individuals labeled as “Other” who were excluded from the training step (n = 79, out of 4,119)^114^. Briefly, each cohort data was subset to a list of 5,847 common SNPs under GRCh38^11^ and then merged at the same sites with reference population data to run genotype PCA. A random forest model was trained on the labeled reference population data to predict population label from the first 10 genotype PCs (with 1,000 decision trees). Then, this classifier was used to infer population labels for individuals from each cohort. The population label with the highest predicted probability was used to assign corresponding GIA to each individual. The individuals with AFR label were considered of AFR GIA for downstream rare variant analysis, whereas the individuals with NFE label were considered of EUR GIA.

#### Analysis of rare variation

Most rare variant analyses were run on three offspring groups from the combined GENAA, ASC, SPARK, and SSC cohorts: AFR GIA, EUR GIA, and ALL (all offspring, independently of their GIA; this includes offspring of other non-AFR and non-EUR GIAs). Rare CNV analysis included any autistic and nonautistic GENAA offspring and just the fully phase-able autistic and nonautistic offspring from ASC, SPARK, and SSC, whereas *de novo* SNV/Indel analysis included only the fully phase-able autistic and nonautistic offspring from the four cohorts. Analyses were performed using R^115^ (version 4.3.1).

#### Rare variant burden analysis

Excess of specific rare variant categories in autistic offspring compared to nonautistic offspring was tested for significance through two-sided binomial tests, with probability of the binomial distribution (*p*) equal to number of autistic individuals divided by the total number of individuals considered for analysis. Nominal p-values < 0.05 were considered significant. Excess was quantified as mean variant count (variant rate) in autistic divided by mean variant count (variant rate) in nonautistic and 95% CI for this variant rate ratio statistic was calculated based on binomial tests. Variant counts underlying these tests are reported in Supplementary Table 22,23.

#### Rare variant carrier enrichment analysis

The significant overlap between autistic individuals and carriers of at least one rare variant of specific categories was tested through two-sided Fisher’s exact tests, using all offspring in the analysis, autistic and nonautistic, as background. Nominal p-values < 0.05 were considered significant. Variant carrier counts underlying these tests are reported in Supplementary Table 22. For this analysis, deletions and duplications were first classified as ASD-associated and/or GDA-associated. Then, the groups of rare CNVs not classified as ASD-associated and/or GDA-associated were further filtered as impacting at least one constrained gene (LOEUF < 0.4). A similar filtering step was used for *de novo* PTVs.

#### Disorder gene set enrichment analysis

The same carrier enrichment analysis approach was used to evaluate if rare variants significantly impacted known associated gene sets. The autosomal protein-coding genes from GENCODE v33 were used as background gene set. The known gene sets of interest, relevant to ASD, were defined as follows. ASD genes scored as 1 (“high-confidence” category), 2 (“strong candidate” category), or S (“syndromic” category) were retrieved from the SFARI Gene database (2024 Q3 release) (https://gene.sfari.org/) and grouped into the following gene lists for testing: SFARI 1, SFARI S, and SFARI 1+2+S (keeping the unique genes from the three unified lists). Neurodevelopmental disorder genes (NDD gene set) were obtained from the Developmental Disorders Gene2Phenotype (DDG2P) database (October 22, 2024 release; https://www.ebi.ac.uk/gene2phenotype/)^125,126^ and further selected to be: (1) dominant or X-linked^12^, (2) impacting brain or cognition, or causing multisystem syndrome based on their organ specificity list^12^, and (3) not with limited Gene2Phenotype association confidence. These four gene sets were further filtered to only include genes within the background through gene symbol matching. Similarly, gene symbol was used to obtain just the rare variants impacting background genes for analysis and to annotate them against the four gene sets. In addition, rare CNVs were annotated as impacting a specific gene set if they impacted at least one background gene that was part of the gene set.

Two types of rare variant carrier enrichment analyses were run, for both rare CNVs and *de novo* SNVs/Indels, through two-sided Fisher’s exact tests. The former tested the significant overlap between autistic individuals and carriers of at least one rare variant (within each tested set, rare deletions, rare duplications, *de novo* PTVs, *de novo* missense SNVs, and *de novo* synonymous SNVs) impacting a specific gene set, using autistic and nonautistic carriers of at least one rare variant impacting the background genes as background (Supplementary Table 23). The latter evaluated the significant overlap between AFR GIA individuals and carriers of at least one rare variant (within each tested set, rare deletions, rare duplications, *de novo* PTVs, and *de novo* missense SNVs) impacting a specific gene set, using AFR GIA and EUR GIA autistic carriers of at least one rare variant impacting the background genes as background (Supplementary Table 22). FDR values (controlling for the simultaneous comparisons at the rare CNV or *de novo* SNV/Indel analysis level) < 0.05 were considered significant.

### GENAA rare variant local genetic ancestry analysis

#### Local ancestry inference

Local ancestry predictions obtained from the GENAA genotyping data with RFMix^65,66^ for the ancestry partial PGS analysis were also used to map the GENAA rare variants to their ancestral background of origin. Within the ancestry partial PGS analysis dataset, only offspring individuals with matched genotyping and WES data could be included in this analysis.

As the rare variants were not phased, it was not possible to determine which chromosome copy they were on and therefore, just rare variants with homozygous diploid local ancestry background (AFR/AFR or EUR/EUR) were annotated and analyzed.

Specifically, for each *de novo* SNV/Indel in a certain fully phase-able carrier, if its position was within a genomic window that was homozygous, and not heterozygous, for either AFR or EUR genetic ancestry, it was possible to assign a local ancestry label to the variant. The process for rare CNV local ancestry inference in autistic and nonautistic carriers was similar, but only CNVs that spanned one or more contiguous genomic segments with consistent homogenous homozygous diploid local ancestry could be annotated.

A slightly different approach was used to maximize local genetic ancestry inference for the non GDA-associated rare deletions in constrained genes in GENAA and SPARK AFR GIA carriers (see below).

#### Local ancestry analysis

For four rare variant sets of interest from GENAA (rare CNVs, *de novo* SNVs/Indels, gnomAD grpmaxSF 1-5% CNVs, and *de novo* missense SNVs), potential differences in variant occurrence within genomic segments with AFR/AFR or EUR/EUR homozygous diploid local ancestry were evaluated. The 701 offspring individuals and 316 fully phase-able offspring individuals in GENAA, included in rare CNV and *de novo* SNV/Indel analysis respectively, were subset to those (1) with available matched QC passing genotyping data and derived local ancestry predictions and therefore, (2) identified as with AFR, EUR, or AFR-EUR admixed genomes based on their genotyping data. After filtering, 427 offspring individuals and 244 fully phase-able offspring individuals from GENAA were left for analysis. The original rare variant sets were subset to variants with AFR/AFR or EUR/EUR homozygous diploid local ancestry and carried by offspring left for analysis (Supplementary Table 24).

For each analyzed individual the total numbers of genome base pairs with AFR/AFR and EUR/EUR homozygous diploid local ancestries were computed and used to obtain their numbers of variants per Mb for AFR/AFR and EUR/EUR genomic regions. This step allowed to normalize rare variant counts by their carrier proportions of AFR/AFR or EUR/EUR global genetic ancestry. There were 11 out of 427 GENAA offspring individuals and 10 out of 244 fully phase-able GENAA offspring individuals with no EUR/EUR base pairs within their genomes and who, consequently, were not included in analyses using numbers of variants per Mb for EUR/EUR genomic regions. Then, offspring were divided into three groups for analysis: autistic, nonautistic, and all. To assess if the individual’s number of variants per Mb for AFR/AFR genomic regions differed from its expected value based on their proportions of AFR/AFR and EUR/EUR global genetic ancestry, the deviation of their intra-individual difference between numbers of variants per Mb for AFR/AFR and EUR/EUR genomic regions from zero was tested with two-sided one-sample paired t-tests^127^. Per variant set FDR values (controlling for the three simultaneous comparisons within each variant set, one for each offspring group) < 0.05 were considered significant. Again, a slightly different approach was used for the analysis of non GDA-associated rare deletions in constrained genes in GENAA and SPARK AFR GIA carriers (see below).

### Analysis of the non GDA-associated rare deletions in constrained genes identified in AFR GIA carriers from the combined GENAA, ASC, SPARK, and SSC cohorts

#### Local genetic ancestry inference and analysis

For 6 out of the 19 AFR GIA carriers of non GDA-associated rare deletions in constrained genes there were no genotyping data and derived local ancestry predictions. For the remaining 14 deletions from 13 carriers with available genotyping data (from the GENAA and SPARK cohorts), the first step was to check if they spanned one or more contiguous genomic segments with consistent homogenous AFR/AFR or EUR/EUR homozygous local ancestry to annotate them as of AFR or EUR origin. Next, for 7 calls (from 7 carriers) with consistent homogenous AFR/EUR heterozygous local ancestry, the available data from fully phase-able carriers’ parents and other family members was used to manually infer the deletion local ancestry. In particular, 5 of these 7 calls (2 carriers had no parental genetic data available) were identified as inherited from a parent, phased, and assigned the correct chromosome copy local ancestry. This two-step annotation approach left 12 annotated deletions of interest (out of 20) from 11 AFR GIA carriers (out of 19) for analysis, of which 6 were annotated as AFR and 6 as EUR.

To test if these calls were more likely to occur within either AFR or EUR genomic segments, 404 GENAA and 292 SPARK AFR GIA offspring individuals part of the rare CNV analysis for whom local ancestry predictions were available were analyzed. Given the subtle differences in genotyping chip technology and genotype imputation quality between the GENAA and SPARK genotyping datasets, local ancestry predictions were obtained over a slightly different total number of genome base pairs in these two cohorts, with around 0.15% additional annotated genome base pairs in GENAA. Therefore, for the combined cohort analysis on 696 GENAA and SPARK AFR GIA offspring individuals, for each individual the total numbers of genome base pairs with AFR and EUR local ancestries (sums of both local ancestry homozygous and heterozygous sites) were first divided by the total number of genome base pairs covered by the individual’s corresponding cohort local ancestry predictions to obtain comparable local ancestry annotated genome normalized Mb values for AFR and EUR genomic regions across individuals. Then, each individual’s AFR and EUR deletion counts were divided by their corresponding Mb values to normalize them by the individual’s proportions of AFR or EUR global genetic ancestry. All 696 offspring individuals were included in analysis since, with the inclusion of local ancestry heterozygous sites, all of them had non-zero AFR or EUR base pairs. For offspring belonging to the autistic, nonautistic, or all groups, two-sided one-sample paired t-tests were used to evaluate if their intra-individual differences between numbers of variants per local ancestry annotated genome normalized Mb for AFR and EUR genomic regions deviated from zero^127^. Per variant set FDR values (controlling for the three simultaneous comparisons, one for each offspring group) < 0.05 were considered significant.

#### Analysis of the deletion-impacted genes

The 20 deletions of interest (19 unique ones) impacted 85 unique genes for which constraint and association with disorders, specifically ASD, NDD, and developmental disorder (DD), was verified. Constraint and association with ASD (using the SFARI 1+2+S gene list) and/or NDD were derived as already described. The additional DD gene set was obtained from the unfiltered DDG2P database (October 22, 2024 release; https://www.ebi.ac.uk/gene2phenotype/)^125,126^ and gene annotation was performed by gene symbol matching. Disorder associated gene lists were defined by selecting the unique genes from the union of those associated with each disorder of interest. Given that the NDD gene set was a subset of the DD gene set, if a gene was annotated as associated with both NDD and DD (basically, with the same disorder), it was only considered specifically associated with NDD for counting and testing. Supplementary Table 26,27 report the per-deletion relevant and per-gene complete constraint and disorder association annotations obtained and tested.

Enrichment of unique deletion-impacted genes for genes known to be associated with ASD and NDD was assessed through two-sided Fisher’s exact test using the autosomal protein-coding genes from GENCODE v33 as background gene set (the 2 unique deletion-impacted genes and the ASD and NDD genes that could not be gene symbol matched to this background were excluded from analysis). The enrichment analysis was also performed by constraint, using only the LOEUF score annotated background genes. Genes with LOEUF < 0.4 were classified as constrained, whereas genes with LOEUF ≥ 0.4 were defined as non-constrained. FDR values (controlling for the three simultaneous tests) < 0.05 were considered significant. Gene counts underlying these tests are reported in Supplementary Table 28.

Moreover, the 77 out of 85 unique deletion-impacted genes with LOEUF score annotation were used to test if the proportions of ASD, NDD and/or DD associated genes differed between constrained and non-constrained genes through two-sided two-sample Pearson’s chi-squared test. Nominal p-value < 0.05 was considered significant.

Finally, enrichment of unique genes impacted by deletions with EUR background for genes tied to ASD and/or DD was assessed with two-sided Fisher’s exact test, using the 15 unique genes impacted by deletions with either EUR or AFR local ancestry as background (Supplementary Table 28). Nominal p-values < 0.05 were considered significant.

#### Visualization of the deletion loci

The Gviz R package^128^ (v1.34.1) was used to build genome browser-like plots for these deletion loci of interest. The deletion impacted gene models displayed in each plot were obtained using the biomaRt R package^129^ (v2.46.3) as follows. First, all the autosomal protein-coding genes from Ensembl v99, corresponding to the GENCODE v33 annotation used for CNV calling, were selected. Then, for each gene only the gene model for its canonical transcript (the same one used for CNV calling), defined as the one with the longest coding sequence or CDS, was displayed. For each deletion the genomic coordinate window capturing the entire length of the gene models for all its impacted genes was shown.

### Analysis of genes impacted by rare variants from AFR GIA autistic carriers

#### Gene list selection

First, all genes (within the rare variant calling background genes, the 19,012 autosomal protein-coding genes from GENCODE v33) impacted by rare variants found in AFR GIA autistic carriers from the combined four cohorts were identified. MPC score annotated *de novo* missense variants, *de novo* PTVs, and rare deletions and duplications affecting one or more genes were considered, and a total of 2,189 genes were identified as mutated in at least one AFR GIA autistic carrier. Within these, 199 had already been associated with ASD (SFARI 1+2+S) and/or NDD and were chosen as the first gene list to test for gene ontology (GO) term enrichment. Then, genes that were constrained and impacted by *de novo* PTVs in at least one AFR GIA autistic carrier (n = 30) were selected as second list. Finally, a third list consisting of the constrained genes impacted by *de novo* PTVs and/or rare deletions in at least one AFR GIA autistic carrier that had not been associated with ASD (SFARI 1+2+S) and/or NDD yet (n = 37) was obtained.

#### GO term enrichment analysis

GO term enrichment analyses were run through the R^115^ (version 4.1.0) package clusterProfiler^130^ (v4.2.2) using the Gene Ontology and Disease Ontology (DO)^73^ annotation term databases, the “org.Hs.eg.db” genome-wide human annotation database, the autosomal protein-coding genes from GENCODE v33 as background gene list, and default parameters. The enrichment of genes from each tested list annotated with a specific term within the background genes with annotation in the term database was evaluated with one-sided Fisher’s exact test. FDR values < 0.05 were considered significant. Fold enrichment for each term was computed dividing the frequency of term annotated list genes by the frequency of term annotated background genes (Supplementary Table 28,29).

#### Protein-protein interaction (PPI) network analysis

Two programs, DAPPLE (Disease Association Protein-Protein Link Evaluator)^74^ (https://www.genepattern.org/) and STRING^75^ (https://string-db.org/), were used to determine if genes from these lists formed significant PPI networks. The four network statistics measured by DAPPLE based on curated PPI data (number of direct connections, seed protein average direct and indirect binding degrees or numbers of interactions, and average common interactor binding degree to the seed proteins) were tested for their significance through 1,000 within degree node label permutations. On STRING, only functional and physical PPI experimental data with at least medium or high (when possible, as for the first 199-gene list) interaction confidence were used to perform PPI enrichment analyses comparing observed and expected numbers of network edges. PPI enrichment p-values < 0.05 were considered significant.

## Data Availability Statement

DNA for the GENAA cohort can be obtained from the National Institute of Mental Health (NIMH) Repository & Genomics Resource (https://www.nimhgenetics.org/). Genetic and behavioral data for the GENAA cohort can be obtained from National Institute of Mental Health Data Archive (NDA) Collection IDs #1879 (NDA Collection DOI: 10.15154/bjds-qd65), #2035 (NDA Collection DOI: 10.15154/c8p8-vx52), and #2976 (NDA Collection DOI: 10.15154/4snm-k786) (NDA Study #2634; NDA DOI: 10.15154/j87q-cr49). Samples and data for the AGRE cohort can be obtained from Autism Speaks (https://www.autismspeaks.org/agre; https://www.autismspeaks.org/applying-access-agre-data-and-biomaterials). Samples and data for the SPARK and SSC cohorts can be obtained from Simons Foundation Autism Research Initiative (SFARI) Base (https://base.sfari.org). Exome data for the ASC cohort can be obtained from dbGaP study accession phs002502.

## Code Availability Statement

Custom code for the analyses reported in this study is available at https://github.com/dhglab/GENAA_genetic_analysis.

---

This **Supplementary Information** file includes:

Supplementary Fig. 1-6

Legends of Supplementary Table 1-29

List of abbreviations used in manuscript

Description of the GENAA Consortium

**Supplementary Fig. 1.**
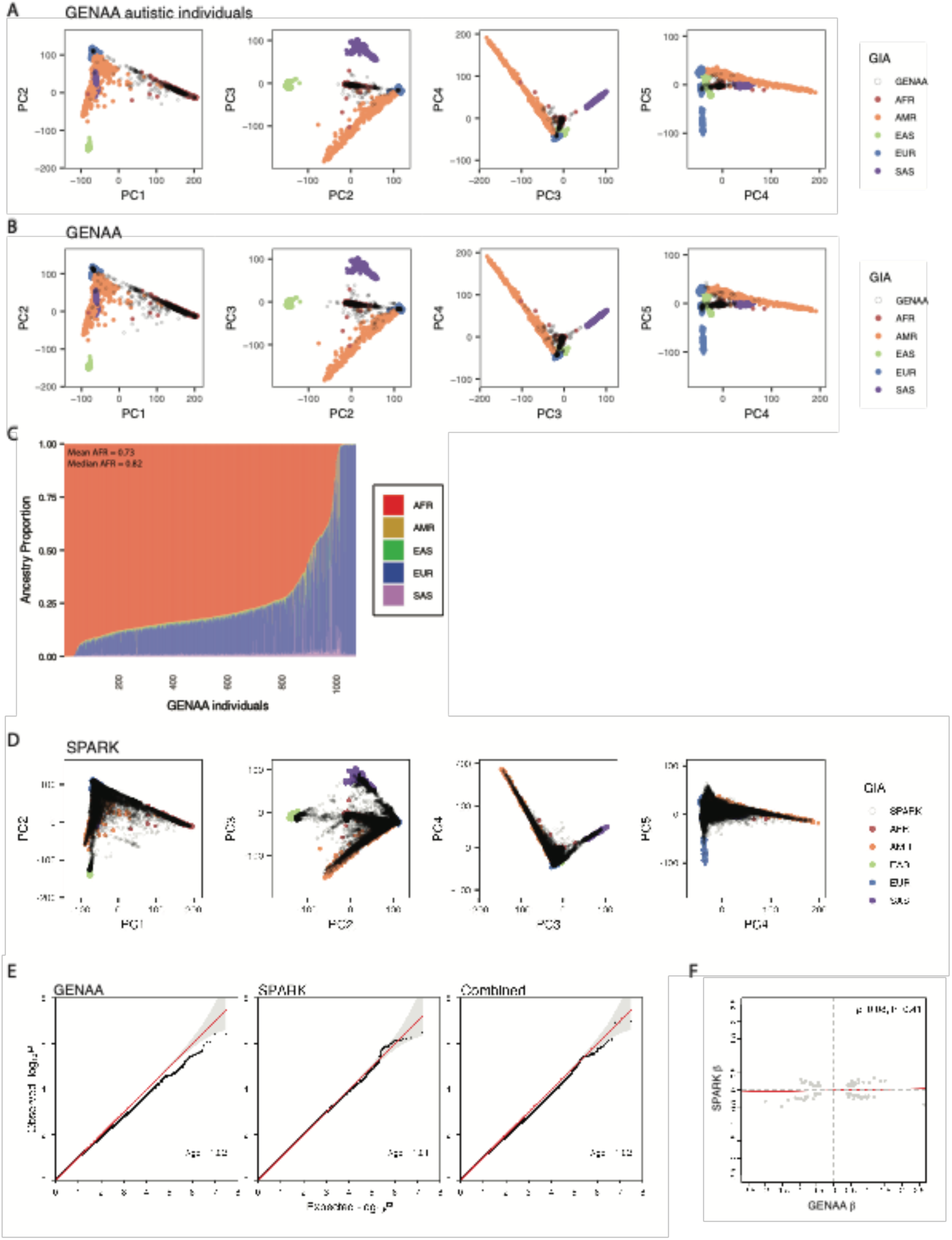
Genetic ancestry prediction in genotyped GENAA and SPARK individuals and AFR GIA based ASD GWAS QC. Refers to Fig. 1. (A-B) Genotype principal component (PC) plots used for GIA prediction, showing the five labeled reference populations’ samples from the 1000 Genomes Project along with the genotyped GENAA (excluding AGRE) autistic individuals (A) and total GENAA (excluding AGRE) individuals (B). Samples from the 1000 Genomes Project are colored by their population label. The GENAA individuals are displayed as empty circles with gray outline and appear overlaid in black on top of their most genetically similar labeled reference population (Methods). (C) Global genetic ancestry proportions estimated for the genotyped GENAA (excluding AGRE) cohort using the five labeled reference populations from the 1000 Genomes Project. Individuals were ordered on the x axis by decreasing AFR global genetic ancestry proportion. Mean and median AFR global genetic ancestry proportions are reported. (D) Genotype PC plots used for GIA prediction of the SPARK individuals used in common variant analysis. The SPARK individuals are shown along with the five labeled reference populations’ samples from the 1000 Genomes Project. Samples from the 1000 Genomes Project are colored by their population label. The SPARK individuals are displayed as empty circles with gray outline and appear overlaid in black on top of their most genetically similar labeled reference population (Methods). (E) Quantile-quantile (Q-Q) plots for ASD diagnosis GWASs on AFR GIA unrelated individuals from GENAA, SPARK, and the two cohorts combined for meta-analysis. These Q-Q plots compare the expected p-values under the null hypothesis of no SNP association on the x axis to the observed p-values on the y axis. The reported λgc (genomic inflation factor) values quantify the degrees of deflation or inflation of the observed p-values, with a value of 1 indicating no deviation from the expected. (F) Spearman correlation of β coefficients of selected SNPs (Methods) from the AFR GIA based ASD GWASs on GENAA unrelated individuals, shown on the x axis, and on SPARK unrelated individuals, reported on the y axis. Top independent SNPs selected for the calculation of the correlation coefficient are described in the Methods.

**Supplementary Fig. 2.**
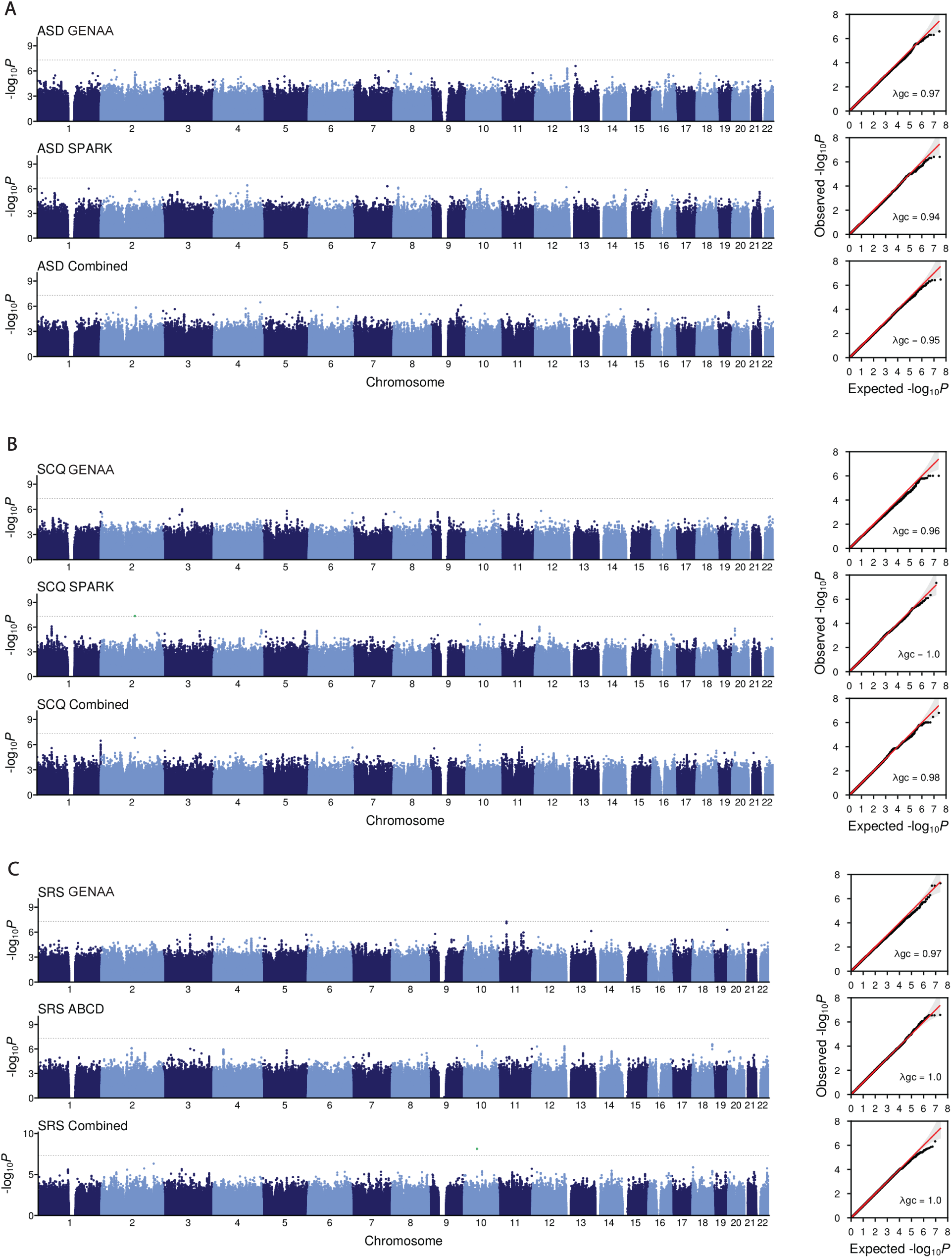
Alternative AFR GIA based GWASs for ASD diagnosis and for quantitative autistic trait measures. Refers to Fig. 1. (A-C) Manhattan plots (left) and quantile-quantile (Q-Q) plots (right) for: (A) ASD diagnosis GWAS on AFR GIA unrelated and related individuals from GENAA, SPARK, and the two cohorts combined for meta-analysis; (B) SCQ raw total score GWAS on AFR GIA unrelated and related individuals from GENAA, SPARK, and the two cohorts combined; (C) SRS raw total score GWAS on AFR GIA unrelated and related individuals from GENAA, ABCD, and the two cohorts combined. The dotted gray horizontal line in each Manhattan plot indicates the genome-wide significance threshold (*P* = 5 x 10^-8^) for SNP association with ASD diagnosis or quantitative autistic trait measure. The reported λgc (genomic inflation factor) values in the Q-Q plots quantify the degrees of deflation or inflation of the observed p-values, with a value of 1 indicating no deviation from the expected p-values under the null hypothesis of no SNP association. Values underlying Supplementary Fig. 2 are reported in Supplementary Table 8-10,12-17.

**Supplementary Fig. 3.**
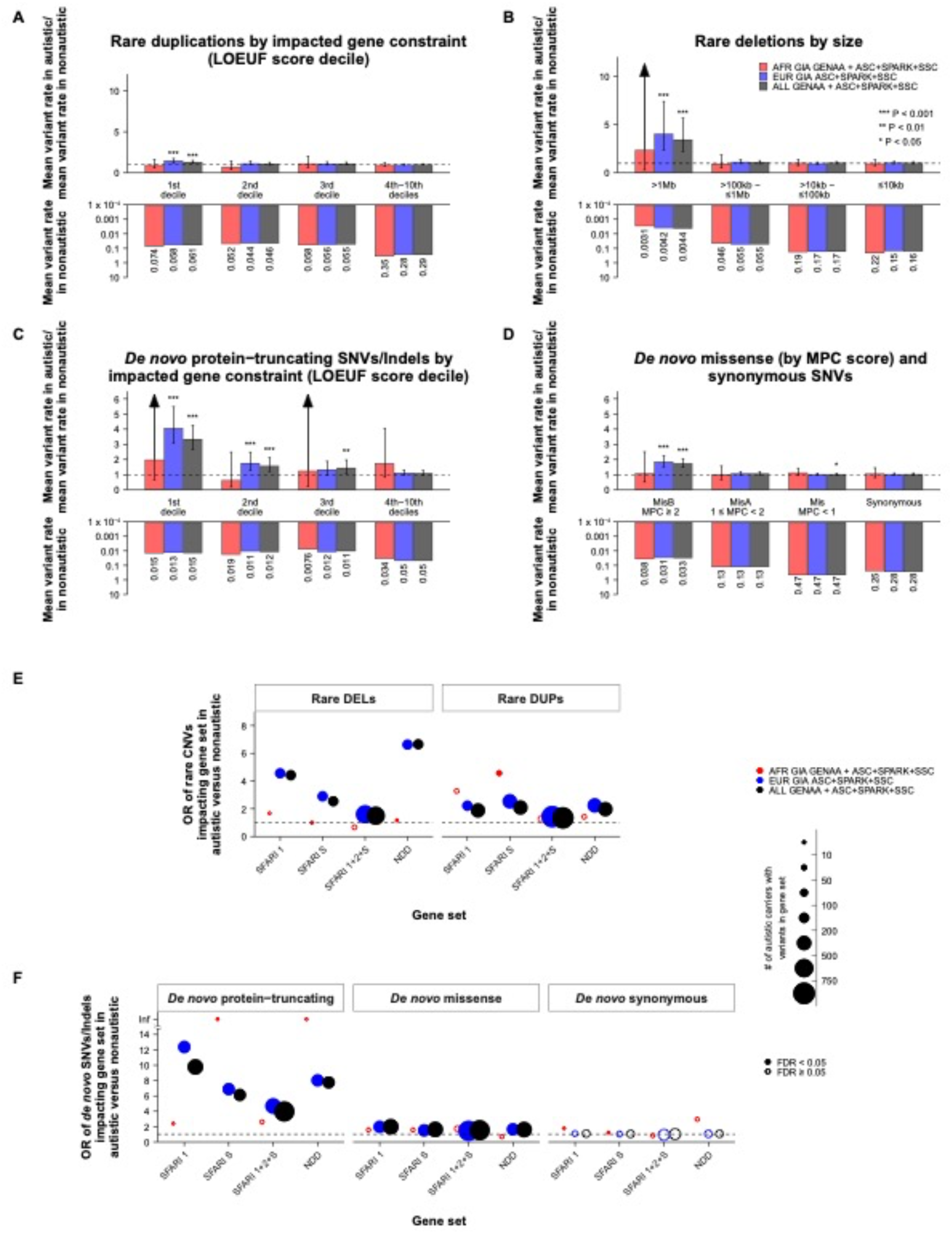
EUR GIA offspring in the ASC, SPARK, and SSC cohorts drive replication of well-known ASD association signals for rare variants and known associated genes in combined analysis with GENAA. Refers to Fig. 3. (A-D) Bar plots showing the autistic by nonautistic mean variant rate ratios (top) and the nonautistic mean variant rates (bottom) for rare duplications by LOEUF score decile of the impacted gene (A), rare deletions by their size category (B), *de novo* PTVs by LOEUF score decile of the impacted gene (C), and *de novo* missense variants by their MPC score along with synonymous variants (D), in three offspring groups based on GIA: AFR (in red), EUR (in blue), and from any GIA (ALL, in black). ASD excess rare variant burden was tested through two-sided one-sample proportion tests via binomial tests on the observed variant counts and proportions of autistic offspring within each tested group. Asterisks show different nominal significance ranges. Error bars represent the 95% CIs for the variant rate ratios, computed through binomial tests. The lower bar plot reversed y axes were log10 transformed for visualization. (E-F) Enrichment of autistic carriers of rare CNVs (deletions or DELs and duplications or DUPs) and *de novo* SNVs/Indels (F) impacting genes belonging to specific gene sets, relevant to ASD (Methods), within each tested group of carriers, AFR, EUR, and ALL. ORs from two-sided Fisher’s exact tests are shown on the y axes. Solid circles represent significant ORs with FDR < 0.05 (per variant set FDR values). Circle size reflects the range for number of autistic carriers of rare CNVs or *de novo* SNVs/Indels impacting each gene set tested. Group sample sizes and test results for analyses underlying Supplementary Fig. 3 are reported in Supplementary Table 23.

**Supplementary Fig. 4.**
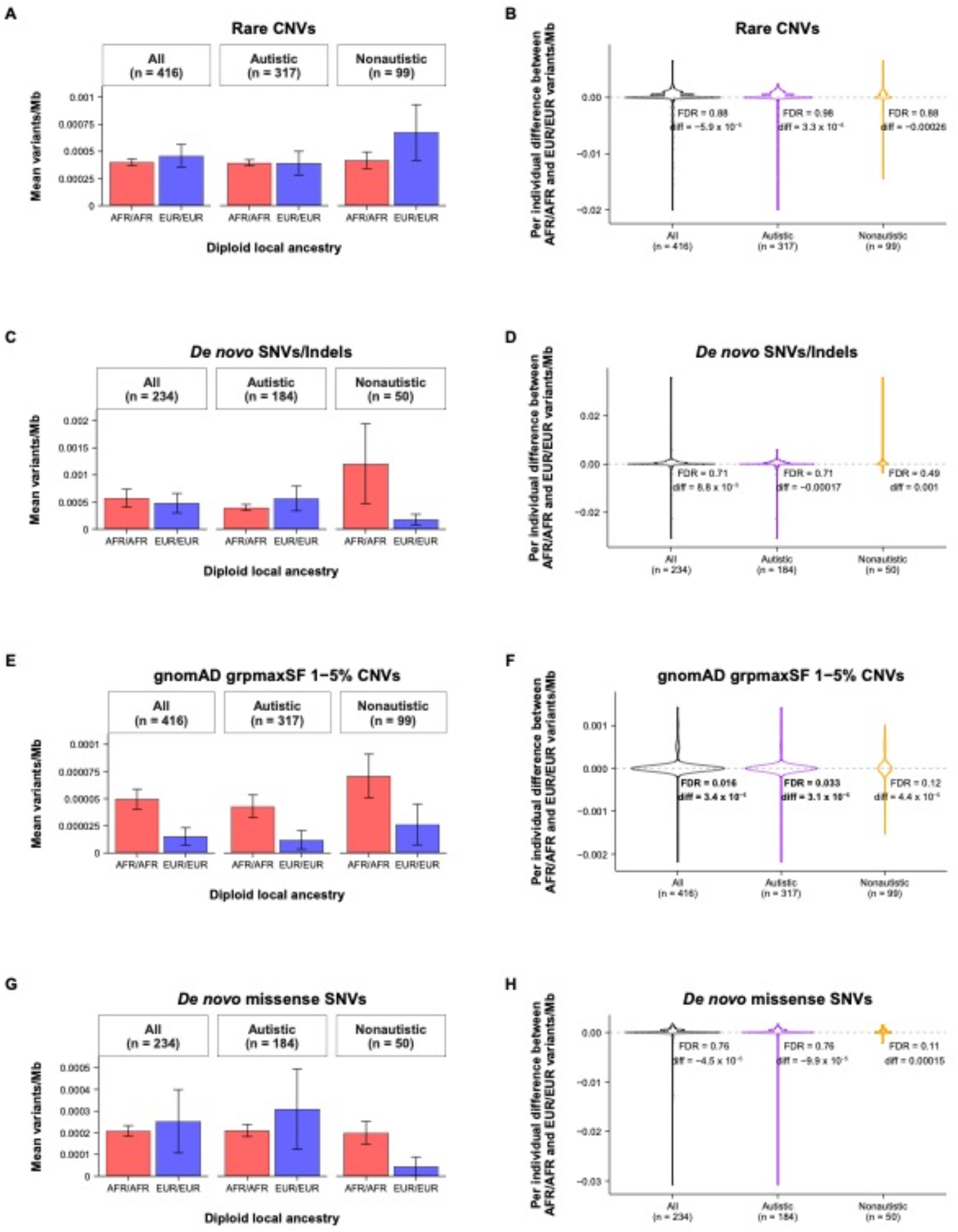
Local genetic ancestry analysis for GENAA rare variant sets of interest: rare CNVs (A-B), *de novo* SNVs/Indels (C-D), gnomAD grpmaxSF 1-5% CNVs (E-F), and *de novo* missense SNVs (G-H). Refers to Fig. 3. (A,C,E,G) Bar plots showing the mean numbers of variants per Mb for genomic regions with AFR/AFR (red) or EUR/EUR (blue) homozygous diploid local ancestry in three GENAA offspring groups with inferred local genetic ancestry (Methods): all, autistic, and nonautistic (further subset to those who are fully phase-able for the *de novo* variant sets). Error bars represent the mean ± standard error values. (B,D,F,H) Violin plots of the distributions of intra-individual differences between numbers of variants per Mb for genomic regions with AFR/AFR homozygous diploid local ancestry and those with EUR/EUR ancestry in three GENAA offspring groups with inferred local genetic ancestry (Methods): all (black), autistic (purple), and nonautistic (orange). Individuals were further subset to those who are fully phase-able for the *de novo* variant set analyses. Per variant set FDR values (controlling for the three simultaneous comparisons within each variant set) from two-sided one-sample paired t-tests are reported together with the mean AFR/AFR minus EUR/EUR differences in variants per Mb (diff). Bold font highlights significant differences with per variant set FDR < 0.05. Values underlying these analyses are reported in Supplementary Table 24.

**Supplementary Fig. 5.**
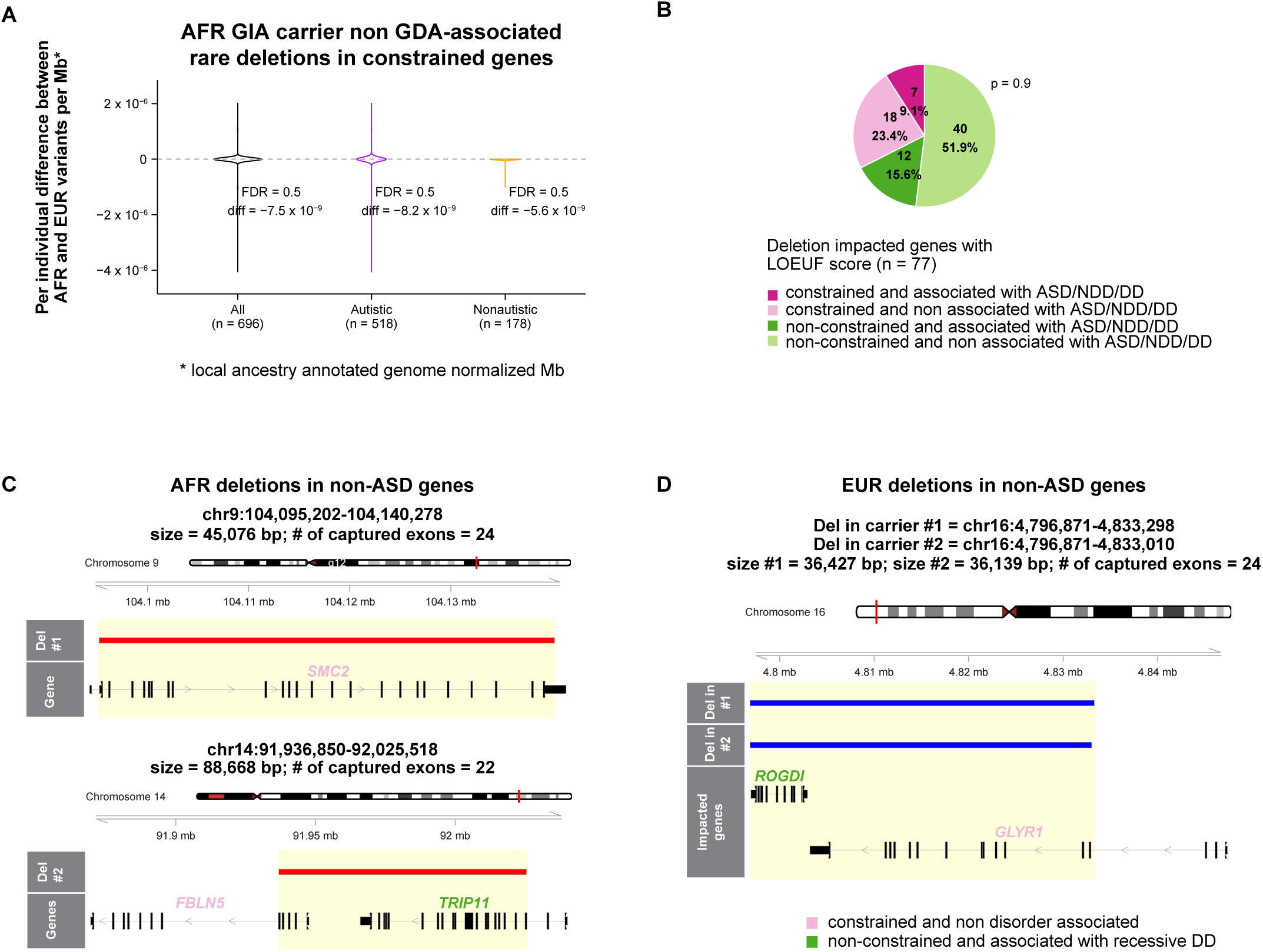
Additional local genetic ancestry and impacted gene analysis of the potentially novel non GDA-associated rare deletions in constrained genes identified in AFR GIA carriers from the combined GENAA, ASC, SPARK, and SSC cohorts. Refers to Fig. 4. (A) Violin plot of the distributions of intra-individual differences between numbers of variants per local ancestry annotated genome normalized Mb for genomic regions with AFR (both homozygous and heterozygous) local ancestry and those with EUR ancestry in three groups of combined GENAA and SPARK AFR GIA offspring with inferred local genetic ancestry (Methods): all (black), autistic (purple), and nonautistic (orange). Per variant set FDR values (controlling for the three simultaneous comparisons) from two-sided one-sample paired t-tests are reported together with the mean AFR minus EUR differences in variants per local ancestry annotated genome normalized Mb (diff). (B) Pie chart showing the percentages of LOEUF score annotated genes impacted by the AFR GIA carrier deletions from the four combined ASD cohorts that are constrained (pink) or not constrained (green) and, additionally, associated (darker shade of pink/green) or not associated (lighter shade) with ASD, NDD and/or DD (Methods). The difference in proportions of disorder associated deletion-impacted genes between the constrained genes and the non-constrained ones was tested through a two-sided two-sample Pearson’s chi-squared test; p-value from this proportion test is reported. (C-D) Visualization of the genomic regions capturing four deletion events and their corresponding impacted gene(s) for three GENAA AFR GIA autistic carriers (Methods). All the deletions, two with AFR (C) and two matching ones with EUR (D) local ancestry, from the same carrier (C) and from two carriers within the same family (D), respectively, impact constrained genes that have not been associated with ASD yet. In addition, three of the shown deletion events also span two non-constrained genes associated with recessive autosomal DDs, *TRIP11* (C) and *ROGDI* (D) (see Supplementary Table 27 for details on the corresponding recessive autosomal DDs). Deletion genomic coordinates, sizes, and numbers of impacted well-captured exons are shown. Analysis values underlying panel (A) are reported in Supplementary Table 28.

**Supplementary Fig. 6.**
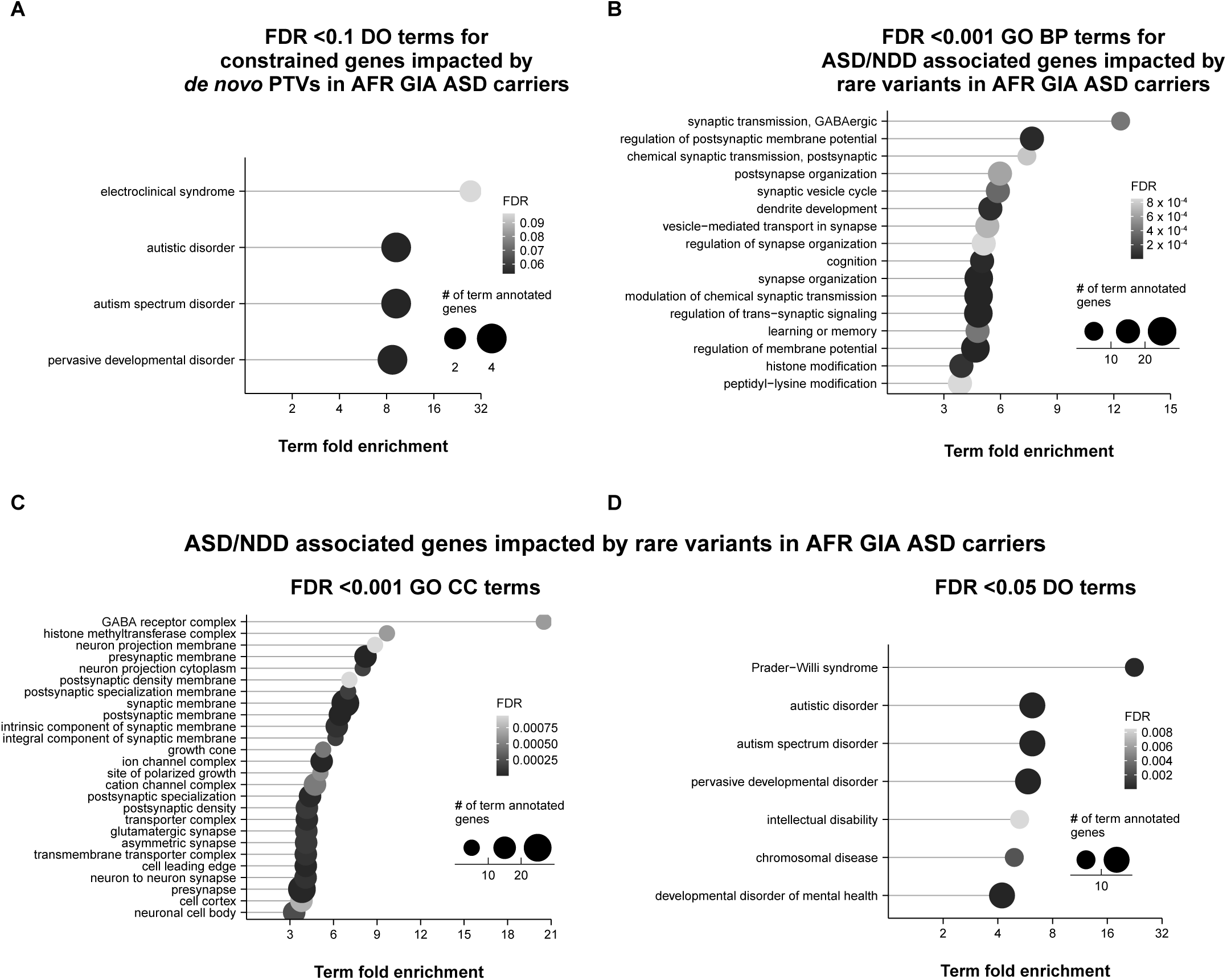
DO and GO term enrichment analyses for the constrained genes impacted by *de novo* PTVs (A) and for the ASD and/or NDD associated genes impacted by rare variants (B-D) in at least one AFR GIA autistic carrier. Refers to Fig. 3 and Fig. 4. (A) Lollipop plot showing the fold enrichment (x axis) (Methods) for DO terms with FDR < 0.1 (y axis) obtained for the constrained genes impacted by *de novo* PTVs in at least one AFR GIA autistic carrier. (B-D) Lollipop plots showing the fold enrichment of top significant (FDR < 0.001) GO biological process (BP) (B) and cellular component (CC) (C) terms and of significant DO (D) terms for the list of disorder associated genes harboring rare variants in at least one AFR GIA autistic carrier. The dot size corresponds to the number of list genes annotated with each specific term, whereas the dot color reflects the one-sided Fisher’s exact test FDR value (Methods). The x axes in panels A,D were log2 transformed for visualization. Analysis values underlying Supplementary Fig. 6 are reported in Supplementary Table 29.

## Legends of Supplementary Tables

**Supplementary Table 1.**

Breakdown of GENAA individuals and families with genotyping and WES data before and after QC.

**Supplementary Table 2.**

Sample manifest for GENAA individuals with genotyping and/or WES data passing QC. The table contains various annotations, including phenotype, GIA label in common and rare variant analyses, respectively, and membership for the AGRE cohort. A total of 2,668 individuals is reported: 1,371 with only genotyping data, 537 with only WES data, and 760 with matched genotyping and WES data.

**Supplementary Table 3.**

Breakdown of GENAA and SPARK individuals with AFR GIA included in common variant analyses. Total numbers of individuals, autistic and nonautistic offspring, and family structures are reported. For offspring, counts by sex and distribution values for age in years are listed. In SPARK, age was recorder at time of enrollment (not at time of diagnosis).

**Supplementary Table 4.**

Sample sizes underlying the different common variant analyses. Counts of individuals from GENAA, SPARK, and ABCD are stratified by phenotype and GIA.

**Supplementary Tables 5-7.**

Top 100 SNP associations from AFR GIA based ASD GWASs with PLINK for GENAA (ST5), SPARK (ST6), and the two cohorts combined (ST7).

**Supplementary Tables 8-10.**

Top 100 SNP associations from AFR GIA based ASD GWASs with SAIGE for GENAA (ST8), SPARK (ST9), and the two cohorts combined (ST10).

**Supplementary Table 11.**

Behavioral assessments for the GENAA study participants.

**Supplementary Tables 12-14.**

Top 100 SNP associations from AFR GIA based SCQ raw total score GWASs for GENAA (ST12), SPARK (ST13), and the two cohorts combined (ST14).

**Supplementary Table 15-17.**

Top 100 SNP associations from AFR GIA based SRS raw total score GWASs for GENAA (ST15), ABCD (ST16), and the two cohorts combined (ST17).

**Supplementary Table 18.**

Local ancestry deconvolution of common SNP rs1452075 (ASD-associated in the EUR-based ASD GWAS) in GENAA and SPARK individuals and autistic offspring from the AFR GIA based ASD GWASs. The table lists observed counts of individuals with the three different genotypes for effect allele T of SNP rs1452075 on AFR background and observed and expected counts of individuals with AFR/EUR, AFR/AFR, or EUR/EUR diploid local ancestry at the SNP locus. P-values from Chi-squared tests are reported.

**Supplementary Table 19.**

Values underlying analyses shown in Fig. 1G-I.

**Supplementary Table 20.**

Values underlying analyses shown in Fig. 2.

**Supplementary Table 21.**

Sample sizes for offspring from GENAA and the other ASD cohorts, ASC, SPARK, and SSC, grouped as AFR, EUR, and ALL, used for rare CNV and *de novo* SNV/Indel analyses.

**Supplementary Table 22.**

Values underlying analyses shown in Fig. 3.

**Supplementary Table 23.**

Values underlying analyses shown in Supplementary Fig. 3.

**Supplementary Table 24.**

Values underlying analyses shown in Supplementary Fig. 4.

**Supplementary Table 25.**

List of GDA-associated and/or ASD-associated CNV events used for rare CNV annotation.

**Supplementary Table 26.**

List of the 20 non GDA-associated rare deletions in constrained genes in 19 AFR GIA carriers, including annotations on deletion local ancestry, impacted well-captured exons, impacted gene constraint, and impacted gene association with ASD, NDD, and/or DD.

**Supplementary Table 27.**

List of 85 unique genes impacted by the non GDA-associated rare deletions in constrained genes in AFR GIA carriers, annotated for their constraint and association with ASD, NDD, and/or DD.

**Supplementary Table 28.**

Values underlying analyses shown in Fig. 4B,C,F,G and Supplementary Fig. 5A.

**Supplementary Table 29.**

Values underlying analyses shown in Supplementary Fig. 6.

### List of abbreviations used in manuscript

ACE: Autism Center of Excellence
ADI-R: Autism Diagnostic Interview-Revised
ADOS-2: Autism Diagnostic Observation Schedule 2
AFR: African
AfrAm: African American
AGRE: Autism Genetic Resource Exchange
AMR: Admixed American
ASC: Autism Sequencing Consortium
ASD: autism spectrum disorder
BP: biological process
CC: cellular component
CI: confidence interval
CNV: copy number variant
DAPPLE: Disease Association Protein-Protein Link Evaluator
DAS-II: Differential Ability Scales-II
DD: developmental disorder
DDG2P: Developmental Disorders Gene2Phenotype
DEL: deletion
DO: disease ontology
DSM-5: Diagnostic and Statistical Manual of Mental Disorders, 5^th^ edition
DUP: duplication
EAS: East Asian
EUR: European
FIN: Finnish
GATK: Genome Analysis Toolkit
GD: genomic disorder
GDA-associated: genomic disorder and/or autism-associated
GENAA: Genetics of Neurodevelopment in African Americans
GIA: genetically inferred ancestry
GIA: genetically inferred ancestry
GLM: generalized linear model
gnomAD: Genome Aggregation Database
GO: gene ontology
GWAS: genome-wide association study
HWE: Hardy-Weinberg equilibrium
IBD: identity-by-descent
LD: linkage disequilibrium
LOEUF: loss-of-function observed/expected upper bound fraction
MAF: minor allele frequency
MID: Middle Eastern
MPC: Missense badness, PolyPhen-2, and Constraint
NDA: National Institute of Mental Health Data Archive
NDD: neurodevelopmental disorder
NFE: Non-Finnish European
NIMH: National Institute of Mental Health
OR: odds ratio
OTH: Other
PC: principal component
PCA: principal component analysis
PGS: polygenic score
PPI: protein-protein interaction
pTDT: polygenic transmission disequilibrium test
PTV: protein-truncating variant
Q-Q plot: quantile-quantile plot
QC: quality control
QS: quality score
SAS: South Asian
SCQ: Social Communication Questionnaire
SE: standard error
SF: site frequency
SFARI: Simons Foundation Autism Research Initiative
SNP: single nucleotide polymorphism
SNV/Indel: single nucleotide variant/small indel
SPARK: Simons Foundation Powering Autism Research for Knowledge
SRS: Social Responsiveness Scale
SSC: Simons Simplex Collection
UNGC: UCLA NeuroGenomics Core
WARP: WDL Analysis Research Pipeline
WDL: Workflow Description Language
WES: whole-exome sequencing

## The Genetics of Neurodevelopment in African Americans (GENAA) Consortium

List of members by their contribution, site, and role:

**Overall study PI:** Daniel H. Geschwind (*University of California Los Angeles*)

**Clinical ascertainment and characterization**

*Emory University*

John N. Constantino (site PI) Ami Klin (site PI)

Cheryl Klaiman (site PI)

*Albert Einstein College of Medicine*

Sophie Molholm (site PI)

*Washington University*

Natasha Marrus (site PI)

*University of California Los Angeles*

Amanda C. Gulsrud (director clinician evaluation)

Aaron D. Besterman (physical and neurological examination) Sunil Mehta (physical and neurological examination)

Tarik Hadzic (physical and neurological examination) Rujuta B. Wilson (physical and neurological examination) Tashalee R. Brown (physical and neurological examination)

*Autism Speaks and Stanford University*

Janet Miller (clinician)

**Biomaterials and sample preparation**

*University of California Los Angeles* Jennifer K. Lowe (project scientist) Hailey Choi (genomic technician)

Jackson N. Hoekstra (genomic technician)

**Study coordination**

*University of California Los Angeles* Jennifer K. Lowe (project scientist) Erin T. Graham (study coordinator)

Marshel F. Adams (study coordinator)

*Washington University*

Anna M. Abbacchi (study coordinator) Yi Zhang (study coordinator)

Amaris N. Clay (study coordinator)

*Albert Einstein College of Medicine*

Elizabeth O. Akinyemi (study coordinator)

**Genetic analysis**

*University of California Los Angeles* Daniel H. Geschwind (site PI) Rita M. Cantor (investigator)

Leanna M. Hernandez (investigator)

Matilde Cirnigliaro (genomic analyst) Michi E. Kumagai (genomic analyst) David S. Gibson (genomic analyst) Jennifer K. Lowe (project scientist)

Stephanie A. Arteaga (genomic technician) Ryan M. Kochis (genomic technician)

Jorge E. Peña-Velasco (genomic technician) Jackson N. Hoekstra (genomic technician)

*University of Pennsylvania*

Bogdan Pasaniuc (site PI)

Alexander O. Flynn-Carroll (genomic analyst)

Kangcheng Hou (genomic analyst; *University of California Los Angeles*)

*Johns Hopkins University*

Dan E. Arking (site PI)

Vamsee Pillalamarri (genomic analyst/technician)

*University of Oxford and University of California San Francisco*

Stephan J. Sanders (site PI)

Shan Dong (genomic analyst/technician)

*Massachusetts General Hospital, Broad Institute of MIT and Harvard, and Harvard Medical School*

Michael E. Talkowski (site PI) Jack M. Fu (genomic analyst)

*State University at Buffalo*

Jamal B. Williams (genomic analyst)

**Manuscript preparation and drafting**

*University of California Los Angeles*

Daniel H. Geschwind (site PI)

Matilde Cirnigliaro (genomic analysis)

David S. Gibson (genomic analysis) Michi E. Kumagai (genomic analysis)

Jennifer K. Lowe (study coordination, data reliability, genomic analysis)

*University of Pennsylvania*

Alexander O. Flynn-Carroll (genomic analysis)

## References

1. Shaw, K. A. et al. Prevalence and Early Identification of Autism Spectrum Disorder Among Children Aged 4 and 8 Years - Autism and Developmental Disabilities Monitoring Network, 16 Sites, United States, 2022. Morb. Mortal. Wkly. Rep. Surveill. Summ. Wash. DC 2002 74, 1–22 (2025).

2. Constantino, J. N. et al. Timing of the Diagnosis of Autism in African American Children. Pediatrics 146, e20193629 (2020).

3. Constantino, J. N. et al. Prospects for Leveling the Playing Field for Black Children With Autism. J. Am. Acad. Child Adolesc. Psychiatry 62, 949–952 (2023).

4. Onovbiona, H., Quetsch, L. & Bradley, R. Racial and Practical Barriers to Diagnostic and Treatment Services for Black Families of Autistic Youth: A Mixed-Method Exploration. J. Autism Dev. Disord. 10.1007/s10803-023-06166-5(2023) doi:10.1007/s10803-023-06166-5.

5. Weitlauf, A. S. et al. Screening, Diagnosis, and Intervention for Autism: Experiences of Black and Multiracial Families Seeking Care. J. Autism Dev. Disord. 54, 931–942 (2024).

6. Maenner, M. J. et al. Prevalence and Characteristics of Autism Spectrum Disorder Among Children Aged 8 Years - Autism and Developmental Disabilities Monitoring Network, 11 Sites, United States, 2020. Morb. Mortal. Wkly. Rep. Surveill. Summ. Wash. DC 2002 72, 1–14 (2023).

7. Gaugler, T. et al. Most genetic risk for autism resides with common variation. Nat. Genet. 46, 881–885 (2014).

8. Weiner, D. J. et al. Polygenic transmission disequilibrium confirms that common and rare variation act additively to create risk for autism spectrum disorders. Nat. Genet. 49, 978–985 (2017).

9. Grove, J. et al. Identification of common genetic risk variants for autism spectrum disorder. Nat. Genet. 51, 431–444 (2019).

10. Ruzzo, E. K. et al. Inherited and De Novo Genetic Risk for Autism Impacts Shared Networks. Cell 178, 850–866.e26 (2019).

11. Satterstrom, F. K. et al. Large-Scale Exome Sequencing Study Implicates Both Developmental and Functional Changes in the Neurobiology of Autism. Cell 180, 568–584.e23 (2020).

12. Zhou, X. et al. Integrating de novo and inherited variants in 42,607 autism cases identifies mutations in new moderate-risk genes. Nat. Genet. 54, 1305–1319 (2022).

13. Fu, J. M. et al. Rare coding variation provides insight into the genetic architecture and phenotypic context of autism. Nat. Genet. 54, 1320–1331 (2022).

14. Antaki, D. et al. A phenotypic spectrum of autism is attributable to the combined effects of rare variants, polygenic risk and sex. Nat. Genet. 54, 1284–1292 (2022).

15. Warrier, V. et al. Genetic correlates of phenotypic heterogeneity in autism. Nat. Genet. 54, 1293–1304 (2022).

16. Weiner, D. J. et al. Statistical and functional convergence of common and rare genetic influences on autism at chromosome 16p. Nat. Genet. 54, 1630–1639 (2022).

17. Cirnigliaro, M. et al. The contributions of rare inherited and polygenic risk to ASD in multiplex families. Proc. Natl. Acad. Sci. U. S. A. 120, e2215632120 (2023).

18. Tick, B., Bolton, P., Happé, F., Rutter, M. & Rijsdijk, F. Heritability of autism spectrum disorders: a meta-analysis of twin studies. J. Child Psychol. Psychiatry 57, 585–595 (2016).

19. Sandin, S. et al. The Heritability of Autism Spectrum Disorder. JAMA 318, 1182–1184 (2017).

20. Yip, B. H. K. et al. Heritable Variation, With Little or No Maternal Effect, Accounts for Recurrence Risk to Autism Spectrum Disorder in Sweden. Biol. Psychiatry 83, 589–597 (2018).

21. Bai, D. et al. Association of Genetic and Environmental Factors With Autism in a 5-Country Cohort. JAMA Psychiatry 76, 1035–1043 (2019).

22. Sandin, S. et al. Examining Sex Differences in Autism Heritability. JAMA Psychiatry 81, 673–680 (2024).

23. Klei, L. et al. Common genetic variants, acting additively, are a major source of risk for autism. Mol. Autism 3, 9 (2012).

24. Cross-Disorder Group of the Psychiatric Genomics Consortium et al. Genetic relationship between five psychiatric disorders estimated from genome-wide SNPs. Nat. Genet. 45, 984–994 (2013).

25. Sirugo, G., Williams, S. M. & Tishkoff, S. A. The Missing Diversity in Human Genetic Studies. Cell 177, 26–31 (2019).

26. Fatumo, S. et al. A roadmap to increase diversity in genomic studies. Nat. Med. 28, 243–250 (2022).

27. Mills, M. C. & Rahal, C. The GWAS Diversity Monitor tracks diversity by disease in real time. Nat. Genet. 52, 242–243 (2020).

28. Martin, A. R. et al. Clinical use of current polygenic risk scores may exacerbate health disparities. Nat. Genet. 51, 584–591 (2019).

29. Wang, Y. et al. Theoretical and empirical quantification of the accuracy of polygenic scores in ancestry divergent populations. Nat. Commun. 11, 3865 (2020).

30. Privé, F. et al. Portability of 245 polygenic scores when derived from the UK Biobank and applied to 9 ancestry groups from the same cohort. Am. J. Hum. Genet. 109, 12–23 (2022).

31. Wojcik, G. L. et al. Genetic analyses of diverse populations improves discovery for complex traits. Nature 570, 514–518 (2019).

32. Lennon, N. J. et al. Selection, optimization and validation of ten chronic disease polygenic risk scores for clinical implementation in diverse US populations. Nat. Med. 30, 480–487 (2024).

33. Ding, Y. et al. Polygenic scoring accuracy varies across the genetic ancestry continuum. Nature 618, 774–781 (2023).

34. Hou, K. et al. Calibrated prediction intervals for polygenic scores across diverse contexts. Nat. Genet. 56, 1386–1396 (2024).

35. Kachuri, L. et al. Principles and methods for transferring polygenic risk scores across global populations. Nat. Rev. Genet. 25, 8–25 (2024).

36. 1000 Genomes Project Consortium et al. A global reference for human genetic variation. Nature 526, 68–74 (2015).

37. Venner, E. et al. The frequency of pathogenic variation in the All of Us cohort reveals ancestry-driven disparities. Commun. Biol. 7, 174 (2024).

38. Manrai, A. K. et al. Genetic Misdiagnoses and the Potential for Health Disparities. N. Engl. J. Med. 375, 655–665 (2016).

39. Chen, S. et al. A genomic mutational constraint map using variation in 76,156 human genomes. Nature 625, 92–100 (2024).

40. Han, A. L. et al. Diverse ancestral representation improves genetic intolerance metrics. Nat. Commun. 16, 2648 (2025).

41. Bryc, K., Durand, E. Y., Macpherson, J. M., Reich, D. & Mountain, J. L. The genetic ancestry of African Americans, Latinos, and European Americans across the United States. Am. J. Hum. Genet. 96, 37–53 (2015).

42. Baharian, S. et al. The Great Migration and African-American Genomic Diversity. PLoS Genet. 12, e1006059 (2016).

43. All of Us Research Program Genomics Investigators. Genomic data in the All of Us Research Program. Nature 627, 340–346 (2024).

44. de Menil, V. et al. The NeuroDev Study: Phenotypic and Genetic Characterization of Neurodevelopmental Disorders in Kenya and South Africa. Neuron 101, 15–19 (2019).

45. Kipkemoi, P. et al. Phenotype and genetic analysis of data collected within the first year of NeuroDev. Neuron 111, 2800–2810.e5 (2023).

46. Cerezo, M. et al. The NHGRI-EBI GWAS Catalog: standards for reusability, sustainability and diversity. Nucleic Acids Res. 53, D998–D1005 (2025).

47. Li, Z. & Zhou, X. Towards improved fine-mapping of candidate causal variants. Nat. Rev. Genet. 10.1038/s41576-025-00869-4 (2025) doi:10.1038/s41576-025-00869-4.

48. Graham, S. E. et al. The power of genetic diversity in genome-wide association studies of lipids. Nature 600, 675–679 (2021).

49. Suzuki, K. et al. Genetic drivers of heterogeneity in type 2 diabetes pathophysiology. Nature 627, 347–357 (2024).

50. Jia, G. et al. Genome-wide association analyses of breast cancer in women of African ancestry identify new susceptibility loci and improve risk prediction. Nat. Genet. 56, 819–826 (2024).

51. Johnson, R. et al. Leveraging genomic diversity for discovery in an electronic health record linked biobank: the UCLA ATLAS Community Health Initiative. Genome Med. 14, 104 (2022).

52. Bhatia, G. et al. Genome-wide scan of 29,141 African Americans finds no evidence of directional selection since admixture. Am. J. Hum. Genet. 95, 437–444 (2014).

53. Hinch, A. G. et al. The landscape of recombination in African Americans. Nature 476, 170–175 (2011).

54. Chang, C. C. et al. Second-generation PLINK: rising to the challenge of larger and richer datasets. GigaScience 4, 7 (2015).

55. Zhou, W. et al. Efficiently controlling for case-control imbalance and sample relatedness in large-scale genetic association studies. Nat. Genet. 50, 1335–1341 (2018).

56. Altman, D. G. & Royston, P. The cost of dichotomising continuous variables. BMJ 332, 1080 (2006).

57. Almasy, L. The role of phenotype in gene discovery in the whole genome sequencing era. Hum. Genet. 131, 1533–1540 (2012).

58. Sham, P. C. & Purcell, S. M. Statistical power and significance testing in large-scale genetic studies. Nat. Rev. Genet. 15, 335–346 (2014).

59. Rutter, M., Bailey, A. & Lord, C. The Social Communication Questionnaire: Manual. (Western Psychological Services, Los Angeles, CA, US, 2003).

60. Yang, J., Lee, S. H., Goddard, M. E. & Visscher, P. M. GCTA: a tool for genome-wide complex trait analysis. Am. J. Hum. Genet. 88, 76–82 (2011).

61. Jernigan, T. L., Brown, S. A. & Dowling, G. J. The Adolescent Brain Cognitive Development Study. J. Res. Adolesc. Off. J. Soc. Res. Adolesc. 28, 154–156 (2018).

62. Brown, S. A., Jernigan, T. L. & Dowling, G. J. The adolescent brain cognitive development study. Health Psychol. Off. J. Div. Health Psychol. Am. Psychol. Assoc. 42, 840–841 (2023).

63. Constantino, J. N. & Gruber, C. Social Responsiveness Scale (SRS) Manual. (Western Psychological Services, Los Angeles, CA, US, 2005).

64. Hou, K. et al. Causal effects on complex traits are similar for common variants across segments of different continental ancestries within admixed individuals. Nat. Genet. 55, 549–558 (2023).

65. Maples, B. K., Gravel, S., Kenny, E. E. & Bustamante, C. D. RFMix: A Discriminative Modeling Approach for Rapid and Robust Local-Ancestry Inference. Am. J. Hum. Genet. 93, 278–288 (2013).

66. Hou, K. et al. Admix-kit: an integrated toolkit and pipeline for genetic analyses of admixed populations. Bioinforma. Oxf. Engl. 40, btae148 (2024).

67. Lambert, S. A. et al. The Polygenic Score Catalog as an open database for reproducibility and systematic evaluation. Nat. Genet. 53, 420–425 (2021).

68. Lambert, S. A. et al. Enhancing the Polygenic Score Catalog with tools for score calculation and ancestry normalization. Nat. Genet. 56, 1989–1994 (2024).

69. Karczewski, K. J. et al. The mutational constraint spectrum quantified from variation in 141,456 humans. Nature 581, 434–443 (2020).

70. Samocha, K. E. et al. Regional missense constraint improves variant deleteriousness prediction. 148353 Preprint at 10.1101/148353 (2017).

71. Gandal, M. J. et al. Broad transcriptomic dysregulation occurs across the cerebral cortex in ASD. Nature 611, 532–539 (2022).

72. Wamsley, B. et al. Molecular cascades and cell type-specific signatures in ASD revealed by single-cell genomics. Science 384, eadh2602 (2024).

73. Yu, G., Wang, L.-G., Yan, G.-R. & He, Q.-Y. DOSE: an R/Bioconductor package for disease ontology semantic and enrichment analysis. Bioinforma. Oxf. Engl. 31, 608–609 (2015).

74. Rossin, E. J. et al. Proteins encoded in genomic regions associated with immune-mediated disease physically interact and suggest underlying biology. PLoS Genet. 7, e1001273 (2011).

75. Szklarczyk, D. et al. The STRING database in 2023: protein-protein association networks and functional enrichment analyses for any sequenced genome of interest. Nucleic Acids Res. 51, D638–D646 (2023).

76. Jurgens, S. J. et al. Rare coding variant analysis for human diseases across biobanks and ancestries. Nat. Genet. 56, 1811–1820 (2024).

77. Mory, A. et al. A nonsense mutation in the human homolog of Drosophila rogdi causes Kohlschutter-Tonz syndrome. Am. J. Hum. Genet. 90, 708–714 (2012).

78. Schossig, A. et al. Mutations in ROGDI Cause Kohlschütter-Tönz Syndrome. Am. J. Hum. Genet. 90, 701–707 (2012).

79. Hirano, T. Condensin-Based Chromosome Organization from Bacteria to Vertebrates. Cell 164, 847–857 (2016).

80. Martin, C.-A. et al. Mutations in genes encoding condensin complex proteins cause microcephaly through decatenation failure at mitosis. Genes Dev. 30, 2158–2172 (2016).

81. Khan, T. N. et al. Mutations in NCAPG2 Cause a Severe Neurodevelopmental Syndrome that Expands the Phenotypic Spectrum of Condensinopathies. Am. J. Hum. Genet. 104, 94–111 (2019).

82. Auer-Grumbach, M. et al. Fibulin-5 mutations link inherited neuropathies, age-related macular degeneration and hyperelastic skin. Brain J. Neurol. 134, 1839–1852 (2011).

83. Safka Brozkova, D., et al. Demyelinating Charcot-Marie-Tooth neuropathy associated with FBLN5 mutations. Eur. J. Neurol. 27, 2568–2574 (2020).

84. Rana, A. P., Ruff, P., Maalouf, G. J., Speicher, D. W. & Chishti, A. H. Cloning of human erythroid dematin reveals another member of the villin family. Proc. Natl. Acad. Sci. U. S. A. 90, 6651–6655 (1993).

85. Khanna, R. et al. Headpiece domain of dematin is required for the stability of the erythrocyte membrane. Proc. Natl. Acad. Sci. U. S. A. 99, 6637–6642 (2002).

86. Ramaswami, G. et al. Integrative genomics identifies a convergent molecular subtype that links epigenomic with transcriptomic differences in autism. Nat. Commun. 11, 4873 (2020).

87. Hampe, W., Rezgaoui, M., Hermans-Borgmeyer, I. & Schaller, H. C. The genes for the human VPS10 domain-containing receptors are large and contain many small exons. Hum. Genet. 108, 529–536 (2001).

88. Savas, J. N. et al. The Sorting Receptor SorCS1 Regulates Trafficking of Neurexin and AMPA Receptors. Neuron 87, 764–780 (2015).

89. De Rubeis, S. et al. Synaptic, transcriptional and chromatin genes disrupted in autism. Nature 515, 209–215 (2014).

90. Zhou, K., Gheybi, K., Soh, P. X. Y. & Hayes, V. M. Evaluating variant pathogenicity prediction tools to establish African inclusive guidelines for germline genetic testing. Commun. Med. 5, 157 (2025).

91. Pathak, A. K. et al. Pervasive ancestry bias in variant effect predictors. 2024.05.20.594987 Preprint at 10.1101/2024.05.20.594987 (2025).

92. Haas, R. et al. Diverse Genomes, Shared Health: Insights from a Health System Biobank. MedRxiv Prepr. Serv. Health Sci. 2025.06.11.25329386 (2025) doi:10.1101/2025.06.11.25329386.

## Methods-only References

93. Robins, D. L. et al. Validation of the modified checklist for Autism in toddlers, revised with follow-up (M-CHAT-R/F). Pediatrics 133, 37–45 (2014).

94. Elliott, C. D. Differential Ability Scales – Second Edition: Administration and Scoring Manual. (Harcourt Assessment, Inc, San Antonio, TX, 2007).

95. Raven, J., Raven, J. C. & Court, J. H. Manual for Raven’s Progressive Matrices and Vocabulary Scales. (Harcourt, San Antonio, TX, 2003).

96. Dunn, L. M. & Dunn, D. M. Peabody Picture Vocabulary Test ( 4th Ed.). (Pearson, 2007).

97. Mullen, E. M. Mullen Scales of Early Learning (AGS Ed.). (American Guidance Service Inc, Circle Pines, MN, US, 1995).

98. Marrus, N. et al. Rapid video-referenced ratings of reciprocal social behavior in toddlers: a twin study. J. Child Psychol. Psychiatry 56, 1338–1346 (2015).

99. Oldfield, R. C. The assessment and analysis of handedness: the Edinburgh inventory. Neuropsychologia 9, 97–113 (1971).

100. Lord, C. et al. Autism Diagnostic Observation Schedule, Second Edition. (Western Psychological Services, Torrance, CA, US, 2012).

101. Lord, C., Luyster, R., Gotham, K. & Guthrie, W. Autism Diagnostic Observation Schedule. 2nd. (ADOS-2) Manual (Part II): Toddler Module. (Western Psychological Services, Torrance, CA, US, 2012).

102. American Psychiatric Association. Diagnostic and Statistical Manual of Mental Disorders: DSM-5. (American Psychiatric Association, Arlington, VA, 2013).

103. Rutter, M., Le Couteur, A. & Lord, C. Autism Diagnostic Interview-Revised. (Western Psychological Services, Los Angeles, CA, US, 2003).

104. Sparrow, S. S., Cicchetti, D. V. & Saulnier, C. A. Vineland Adaptive Behavior Scales, Third Edition (Vineland-3). (Pearson, San Antonio, TX, US, 2016).

105. Achenbach, T. M. Manual for the Child Behavior Checklist/4-18 and 1991 Profile. (University of Vermont, Department of Psychiatry, Burlington, VT, 1991).

106. Sanchez, M. J. & Constantino, J. N. Expediting clinician assessment in the diagnosis of autism spectrum disorder. Dev. Med. Child Neurol. 62, 806–812 (2020).

107. Lajonchere, C. M. & AGRE Consortium. Changing the landscape of autism research: the autism genetic resource exchange. Neuron 68, 187–191 (2010).

108. Taliun, D. et al. Sequencing of 53,831 diverse genomes from the NHLBI TOPMed Program. Nature 590, 290–299 (2021).

109. Loh, P.-R. et al. Reference-based phasing using the Haplotype Reference Consortium panel. Nat. Genet. 48, 1443–1448 (2016).

110. Manichaikul, A. et al. Robust relationship inference in genome-wide association studies. Bioinformatics 26, 2867–2873 (2010).

111. SPARK Consortium. SPARK: A US Cohort of 50,000 Families to Accelerate Autism Research. Neuron 97, 488–493 (2018).

112. Reich, D., Price, A. L. & Patterson, N. Principal component analysis of genetic data. Nat. Genet. 40, 491–492 (2008).

113. Fairley, S., Lowy-Gallego, E., Perry, E. & Flicek, P. The International Genome Sample Resource (IGSR) collection of open human genomic variation resources. Nucleic Acids Res. 48, D941–D947 (2020).

114. National Academies of Sciences, Engineering, and Medicine. Using Population Descriptors in Genetics and Genomics Research: A New Framework for an Evolving Field. (The National Academies Press, Washington, DC, 2023).

115. R Core Team. R: A Language and Environment for Statistical Computing. R Foundation for Statistical Computing (2023).

116. Privé, F., Aschard, H., Ziyatdinov, A. & Blum, M. G. B. Efficient analysis of large-scale genome-wide data with two R packages: bigstatsr and bigsnpr. Bioinforma. Oxf. Engl. 34, 2781–2787 (2018).

117. Willer, C. J., Li, Y. & Abecasis, G. R. METAL: fast and efficient meta-analysis of genomewide association scans. Bioinformatics 26, 2190–2191 (2010).

118. Kim, M., Vo, D. D., Kumagai, M. E., Jops, C. T. & Gandal, M. J. GeneticsMakie.jl: a versatile and scalable toolkit for visualizing locus-level genetic and genomic data. Bioinformatics 39, btac786 (2023).

119. Voss, K., Gentry, J. & Auwera, G. V. der. Full-stack genomics pipelining with GATK4 + WDL + Cromwell [version 1; not peer reviewed]. F1000Research 6, 1379 (poster) (2017).

120. Degatano, K. et al. WDL Analysis Research Pipelines: Cloud-Optimized Workflows for Biological Data Processing and Reproducible Analysis. Preprint at 10.20944/preprints202401.2131.v1 (2024).

121. Regier, A. A. et al. Functional equivalence of genome sequencing analysis pipelines enables harmonized variant calling across human genetics projects. Nat. Commun. 9, 4038 (2018).

122. Babadi, M. et al. GATK-gCNV enables the discovery of rare copy number variants from exome sequencing data. Nat. Genet. 55, 1589–1597 (2023).

123. McLaren, W. et al. The Ensembl Variant Effect Predictor. Genome Biol. 17, 122 (2016).

124. Robinson, J. T., Thorvaldsdóttir, H., Wenger, A. M., Zehir, A. & Mesirov, J. P. Variant Review with the Integrative Genomics Viewer. Cancer Res. 77, e31–e34 (2017).

125. Thormann, A. et al. Flexible and scalable diagnostic filtering of genomic variants using G2P with Ensembl VEP. Nat. Commun. 10, 2373 (2019).

126. Yates, T. M. et al. Curating genomic disease-gene relationships with Gene2Phenotype (G2P). Genome Med. 16, 127 (2024).

127. Kessler, M. D. et al. De novo mutations across 1,465 diverse genomes reveal mutational insights and reductions in the Amish founder population. Proc. Natl. Acad. Sci. U. S. A. 117, 2560–2569 (2020).

128. Hahne, F. & Ivanek, R. Visualizing Genomic Data Using Gviz and Bioconductor. in Statistical Genomics: Methods and Protocols (eds. Mathé, E. & Davis, S.) 335–351 (Springer, New York, NY, 2016). doi:10.1007/978-1-4939-3578-9_16.

129. Durinck, S., Spellman, P. T., Birney, E. & Huber, W. Mapping identifiers for the integration of genomic datasets with the R/Bioconductor package biomaRt. Nat. Protoc. 4, 1184–1191 (2009).

130. Yu, G., Wang, L.-G., Han, Y. & He, Q.-Y. clusterProfiler: an R package for comparing biological themes among gene clusters. Omics J. Integr. Biol. 16, 284–287 (2012).

